# HAVOC: Small-scale histomic mapping of biodiversity across entire tumor specimens using deep neural networks

**DOI:** 10.1101/2023.01.11.22283903

**Authors:** Anglin Dent, Kevin Faust, K. H. Brian Lam, Narges Alhangari, Alberto J. Leon, Queenie Tsang, Zaid Saeed Kamil, Andrew Gao, Prodipto Pal, Stephanie Lheureux, Amit Oza, Phedias Diamandis

**Author notes:** Please direct correspondence to: **Phedias Diamandis, MD, PhD, FRCPC**, Neuropathologist, Department of Pathology, University Health Network, 12-308, Toronto Medical Discovery Tower (TMDT), 101 College St, Toronto, M5G 1L7. equal contributions (Anglin Dent, Kevin Faust, Phedias Diamandis).

## Abstract

**Summary:** Intra-tumoral heterogeneity can wreak havoc on current precision medicine strategies due to challenges in sufficient sampling of geographically separated areas of biodiversity distributed across centimeter-scale tumor distances. In particular, modern tissue profiling approaches are still largely designed to only interrogate small tumor fragments; which may constitute a minute and non-representative fraction of the overall neoplasm. To address this gap, we developed a pipeline that leverages deep learning to define topographic histomorphologic fingerprints of tissue and create Histomic Atlases of Variation Of Cancers (HAVOC). Importantly, using a number of spatially-resolved readouts, including mass-spectrometry-based proteomics and immunohistochemisy, we demonstrate that these personalized atlases of histomic variation can define regional cancer boundaries with distinct biological programs. Using larger tumor specimens, we show that HAVOC can map spatial organization of cancer biodiversity spanning tissue coordinates separated by multiple centimeters. By applying this tool to guide profiling of 19 distinct geographic partitions from 6 high-grade gliomas, HAVOC revealed that distinct states of differentiation can often co-exist and be regionally distributed across individual tumors. Finally, to highlight generalizability, we further benchmark HAVOC on additional tumor types and experimental models of heterogeneity. Together, we establish HAVOC as a versatile and accessible tool to generate small-scale maps of tissue heterogeneity and guide regional deployment of molecular resources to relevant and biodiverse tumor niches.

---

Tumoral heterogeneity underpins modern frameworks of tumor evolution and treatment resistance, but resolving cancer biodiversity distributed across long distances in resection specimens has proven challenging^1–3^. Even small biopsies (∼1 cm^3^) can contain ∼10^8^ tumor cells; a number representing many orders of magnitude more than what current single-cell profiling approaches can routinely process (∼10^3^ cells)^4–6^. Similarly, asymmetric distributions of biological variation across specimens can result in non-representative under-sampling and/or mixing of critical tumor subpopulations using bulk-based expression profiling approaches^7,8^. These complexities may be further amplified in large cohort studies and analysis of recurrent tumor specimens where variations in cellular composition across samples can complicate interpretations^9–11^.

In geography, large-scale maps that provide high-resolution information of relatively small topographic areas (e.g. cities, provinces) are often complemented with smaller-scale cartographic surveys that document specific sets of relevant features over larger regions (e.g. countries and continents)^12,13^. In such disciplines, these small-scale atlases aid in guiding systems-level decisions and resource allocation (e.g. nature conservation efforts). In the context of cancer, systematic approaches to establishing coordinates of potential biodiversity across large tissue specimens could guide deployment of limited molecular profiling resources to better capture tumor-level heterogeneity^14^ (**Fig 1a**).

**Figure 1.**
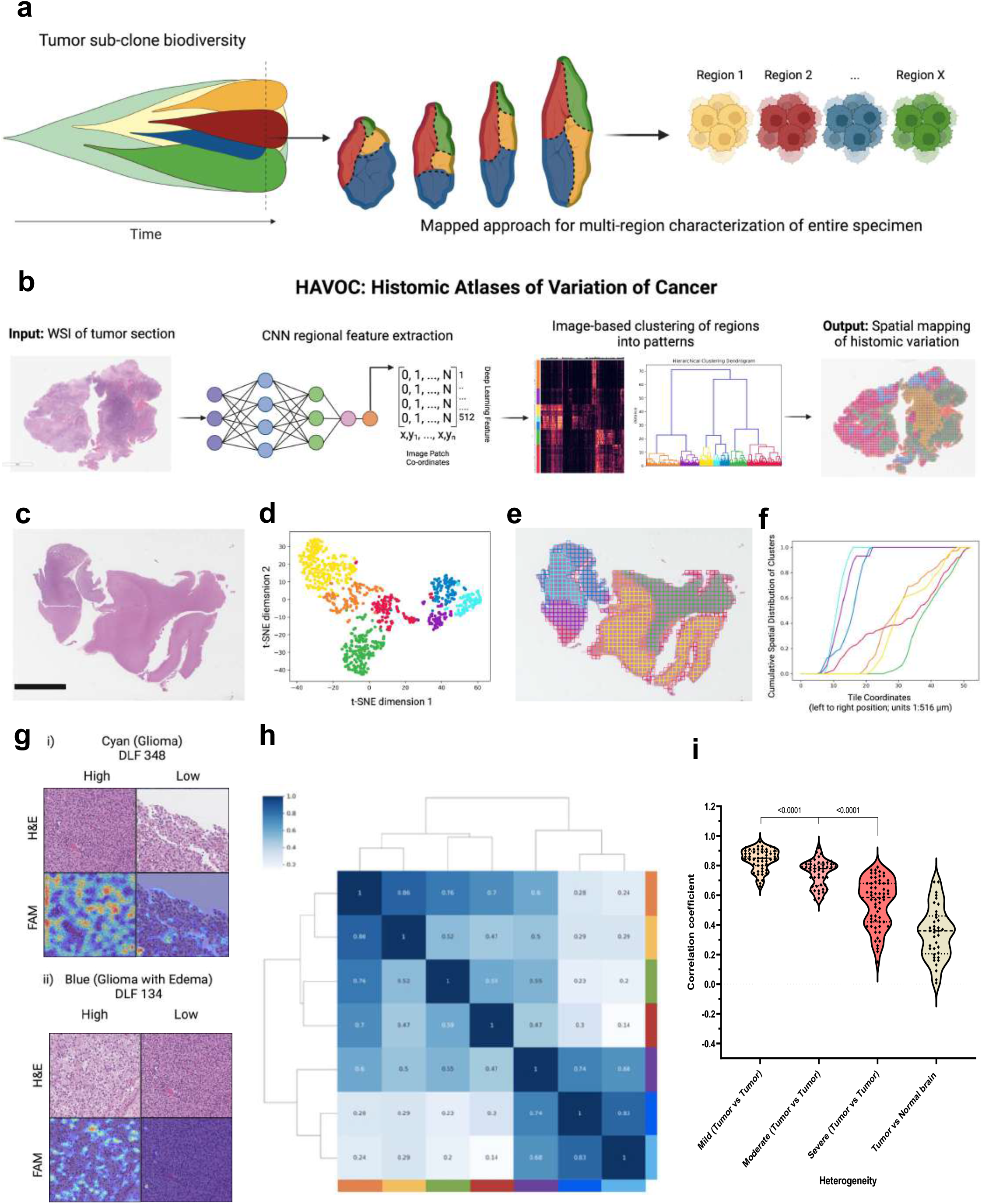
Mapping biodiversity in cancer tissue with HAVOC. **(a)** Cartoon depicting regional evolution of intra-tumoral subclones and a map-guided approach to sampling. **(b)** HAVOC workflow summary. A pre-trained convolutional neural network (CNN) is used as a morphology feature extractor for input images. The generated deep learning feature vectors (DLFVs) of individual tiles are then used to carry out image-based clustering and map spatial coordinates to distinct histomic fingerprints back to the original whole slide image (WSI). **(c-e)** Representative mapping of a diffuse glioma. The relative spectrum of morphologic patterns of image tiles can be explored by dimensionality reduction (e.g. t-SNE) and clustering. Both highlight distinct tumor (blue, purple cyan) and non-tumoral (yellow, green, red) regions. Scale bar: 6 mm. **(f)** Horizontal (vertical left to right raster) cumulative distribution plot of the overall fraction of tiles from each HAVOC-defined cluster in panel d across entire WSI highlighting non-random spatial distributions of HAVOC-defined histomorphologies (Kolmogorov–Smirnov statistic of horizontal distribution of cellular tumorcyan vs cellular tumor with edemablue = 0.69, p=1.2e^-11^) **(g)** Mapping of individual DLFs over-represented in these HAVOC-defined tumor regions highlight interpretable morphological patterns of these defined glioma niches (e.g. tumor nuclei and edema respectively). **(h)** Pairwise Pearson correlation coefficients (*r*) of the DFLVs of HAVOC-defined partitions in panel (e) highlighting inverse correlation with the degree morphological heterogeneity (and t-SNE distances in panel (d)) across this representative case **(i)** Violin plots showing concordance between HAVOC *r* values and semi-quantitative assessments of regional heterogeneity (e.g. mild, moderate, or severe) by experts. DLFV *r* of lesional vs non-lesional regions included as reference.

To address this critical challenge, we developed HAVOC, a histology-based deep neural network pipeline aimed at creating “Histomic Atlases of Variation Of Cancers” and providing spatially contextualized estimates of biodiversity across virtually any scale (**Fig 1b**). This approach leverages both classic and modern dogmas of histopathology; (i) that significant changes in molecular programs are often accompanied with modifications in cellular morphologies, and (ii) that such cytoarchitectural variations can be objectively defined by contemporary computer vision strategies^15–18^. Previous studies exploring intra-tumoral heterogeneity using computer vision have largely used supervised approaches, in which neural networks are trained using hematoxylin and eosin (H&E)-stained whole slide images (WSI) with genetically-defined ground truth labels generated from bulk tumor tissue (e.g. TCGA)^19–21^. Once trained these models are then applied to predict the mutational, cell composition and transcriptional status of individual image patches from larger tissue areas. While such approaches have shown capacity to predict presence of immune infiltrates and a handful of specific mutations in a context-specific manner, more intricate tumor cell-intrinsic biological programs (e.g. proliferation, hypoxia, DNA repair) have proven more challenging to generalize across the majority of tumor types^22–24^. Similarly, such approaches usually require knowledge and design around specific mutations defined *a priori* (e.g. FGFR3 mutations in bladder cancer) and may therefore be less suitable for screening for potential intra-tumoral heterogenous subclones that may be patient-specific and emerge downstream of these common initiating genetic events^25,26^. To address this, one recent study used deep learning to detect immune and stroma infiltrates and leveraged these features as geographic guideposts to define immune “cold” and “hot” regions of non-small cell lung carcinomas. Using whole-exome and RNA-sequencing on these deep-learning defined regions, they showed that cancer subclones derived from immune cold regions shared closer proximity in mutational space than subclones from immune hot regions^14^. Such molecularly-agnostic and prospective mapping approaches of geospatial variability, are critical as they may provide more personalized and precise insights into the emergence of treatment-resistant subclones and aggressive clinical phenotypes^27,28^.

Here, we show that by using unsupervised partitioning of patterns found on H&E-stained WSIs, HAVOC can predict the precise coordinates and provide relative estimates of relevant morphologic and molecular patterns of cancer diversity. Importantly, this approach does not require any *a priori* framework of biodiversity, allowing for tumor heterogeneity to be explored free of any pre-defined constraints (e.g. regional lymphocytic infiltrates) or narrow molecular definitions (e.g. specific mutations, aneuploidy) that may not robustly generalize in molecularly heterogenous cancers. Specifically, using high grade gliomas as a proof-of-concept, an aggressive form of brain cancer that can show significant intra-tumoral variation, HAVOC’s predictions of biovariation showed strong concordance with both human- and molecularly-defined estimates of heterogeneity. This regional analysis also specifically revealed that tumor subpopulations with varying degrees of phenotypic differentiation and proliferative capacity can often co-exist and be geographically separated in high grade gliomas; underscoring the need for more intelligent sampling strategies. This unique spatially-defined glioma proteomic dataset generated in this study also showed additional patient-specific regional differences that can be further explored in real-time in the Brain Protein Atlas^29^ (https://www.brainproteinatlas.org/dash/apps/ad). Importantly, we further highlight the generalizability of HAVOC to other tumor types and cancer models and show it can be deployed without the need for any additional context-specific training. In addition to the available code for local implementation, we developed a server version of HAVOC that can accessed via a web interface (https://www.codido.co) without the need for any specialized hardware or software expertise. Routine generation of such small-scale histomic atlases of biologic variations in cancer offers an opportunity to better identify and document critically divergent cellular habitats across large geographic regions and may aid in better managing tumor heterogeneity for both large cohort studies and personalized medicine efforts.

## Prediction and relative quantification of histomorphological heterogeneity by HAVOC

To assess if delineation and quantification of spatial transitions in histomorphology could be automated, we applied an unsupervised image clustering framework^30^ to a digitized WSI cohort of brain tumors comprising largely of diffuse gliomas (n=40) showing varying degrees of regional heterogeneity. The advantage of this unsupervised deep learning strategy is that it helps overcome the significant case-to-case morphologic variability that is seen in many malignant cancers such as high grade gliomas^31^.

Briefly, WSIs are individually partitioned into 0.066-0.27 mm^2^ non-overlapping image patches (based on specimen size and pattern of interest) and passed through a histology-optimized convolutional neural network (CNN) previously trained on a diverse set of nearly 1 million pathologist-annotated image patches extracted from over 1,000 brain tumors^15,32^. Rather than carrying out classification, the 512-dimensional deep learning feature vector (DLFV), generated in the penultimate layer of the CNN, is extracted and used as a “histomic” feature set. These DLFV signatures are then utilized to carry out patch-level clustering (k=2-9) and spatially map morphologic variation across entire WSIs (**Fig 1c-e, Supplemental Fig 1**). In addition to lesion segmentation from normal brain tissue elements in early partitions, when present, this approach also non-randomly segregated tumor areas with progressively more subtle regional morphologic pattern differences in later subdivisions (e.g. variations in tumor cellularity and intra-tumoral edema, p=1.2e^-11^, Kolmogorov–Smirnov test) (**Fig 1f, Supplemental Fig 2**). Generation of feature activation maps (FAMs) of salient individual deep learning features (DLFs), enriched in each compartment, provides support and insight of human-perceivable histomorphologic patterns associated with each HAVOC partition (**Fig 1g**). Importantly, Pearson correlation coefficients (*r*) of regional DLFVs correlated with human expert estimates of histologic variation; allowing this metric to serve as a quantitative approximation of the degree of variability across HAVOC-defined regions (**Fig 1h-i**, n=40 cases; *p*<0.0001, Mann-Whitney U test). While we found that the optimal number of subdivisions varied depending on the degree of tissue complexity found on individual slide, for most cases, the majority of discernible patterns of histomorphologic variation (*r* = ∼0.74) plateaued and reached saturation after 7-8 partitions; irrespective of the overall level of heterogeneity found on the WSI **(Supplemental Fig 3)**. Together, this data highlights how HAVOC can provide an automated, objective, and human expert-concordant tool to estimate spatial histomorphologic variation across cancer tissue.

## Spatial morphologic fingerprints align with molecular patterns of heterogeneity

Interestingly, in one of the glioblastoma cases of our initial cohort, we noted that HAVOC partitioning captured a BRAFV600E-mutated tumor subclone showing an elevated Ki-67 (MIB1) proliferation index on immunohistochemistry (**Fig 2a**). This supported the possibility that regional changes in morphologic fingerprints may also predict relevant phenotype-level variability in tumor biology. To begin formally testing if histomic heterogeneity, perceived by HAVOC, also correlated with global molecular differences, we compared the *r* of DLFVs across the major histomorphologic hallmarks of diffuse gliomas including regions of high cellularity (CT), infiltrating tumor (IT) and brain tissue at the leading tumor edge (LE) to proteomic and transcriptional profiles generated from these areas in previous studies^33,34^ (**Supplemental Fig 4**). Indeed, HAVOC-defined niche variations correlate with proteomic (r^2^=0.79) and transcriptomic (r^2^=0.89) profiles with similar glioma histomorphologic regions (eg. multiple IT regions) having a higher degree of DLFV and molecular similarity when compared to more distinct niches (eg. CT vs LE regions).

**Figure 2.**
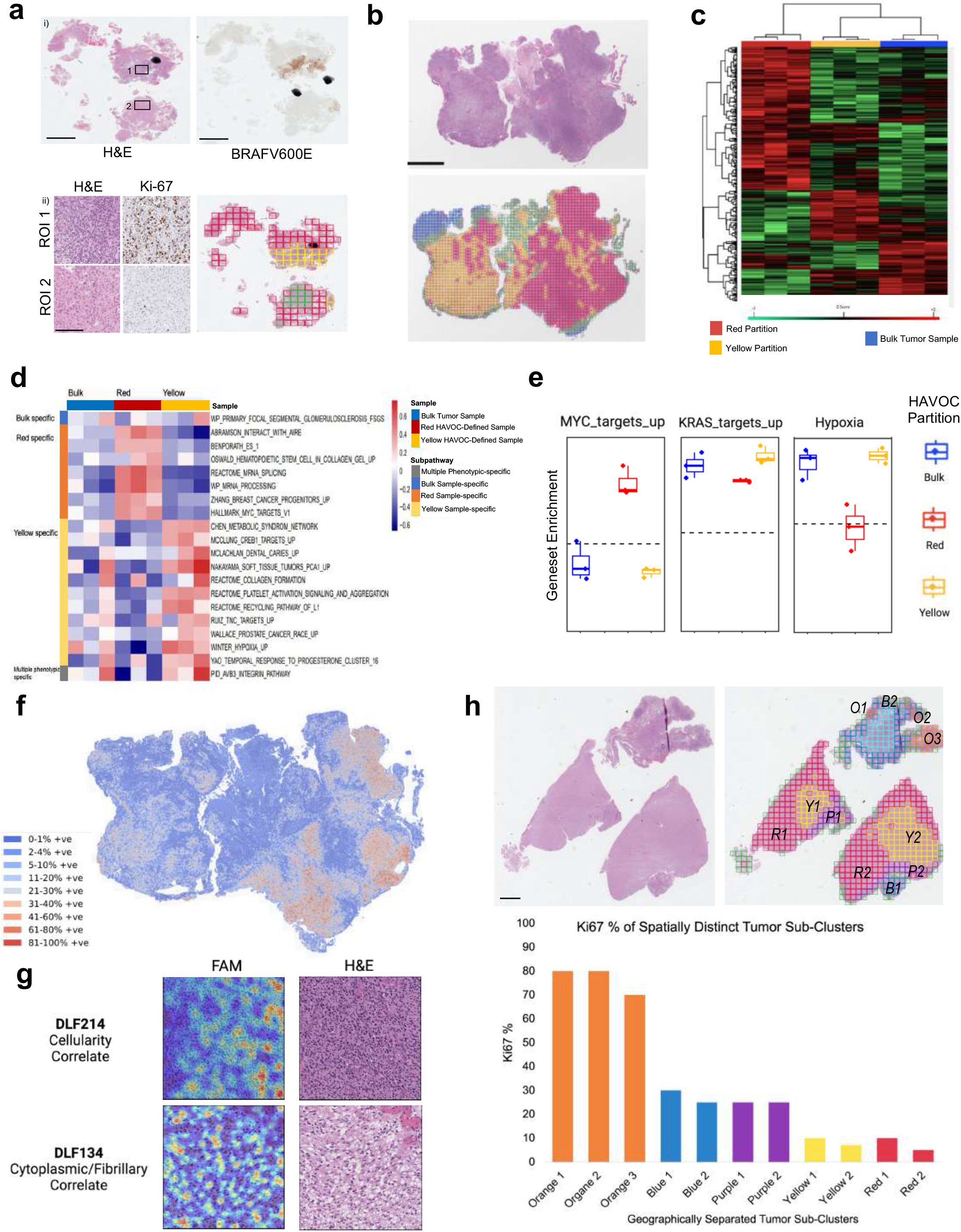
HAVOC-defined partitions align with regional biodiversity. **(a)** Histopathology images of an IDH-wildtype glioblastoma demonstrating intra-tumoral heterogeneity with a BRAFV600E-mutated hyper-proliferative subclone resolved by immunohistochemistry. This genetic and biologically-defined subclone was resolved by a specific HAVOC partition (yellow). Scale bar: 2 mm. **(b)** WSI image of a glioblastoma mapped with HAVOC. Scale bar: 4 mm. **(c)** LC-MS/MS profiling of HAVOC-defined partitions from panel (b) shows distinct molecular profiles from each other and the overall (bulk) specimen. **(d-e)** Single-sample Gene Set Enrichment Analysis highlights heterogeneity across various programs and relevant glioma axes including MYC (proliferation), KRAS (invasion), and hypoxia. Bulk signatures provided for reference. **(f)** Spatial proliferation index differences (as assessed via CNN-based quantification of Ki-67) spatially align with HAVOC partition. (**g**) Feature activation mapping define morphologic correlates of the profiled HAVOC partitions. Partitions showing proliferative and invasive biology show high cellularity and cytoplasmic/fibrillary patterns respectively. **(h)** Representative H&E and HAVOV map of an IDH-wildtype glioblastoma section with geographically separated subclusters (e.g. Orange O1, O2, O3) showing similarly grouped histomic (HAVOC) signatures. Despite distinct spatial coordinates, subclusters belonging to the same HAVOC partitions displayed similar biology (e.g. Ki-67 proliferation indices). Scale bar: 2 mm.

To more directly assess the predictive power of HAVOC for resolving distinct spatial expression profiles, we next determined if regional histomically-defined niches, exclusively within the cellular glioma compartment, could predict phenotypic protein-level heterogeneity. We focused our regional analysis on proteomic outputs due to known proteogenomic discordances in many cancers in which genetic changes may not always translate to downstream cellular phenotypes^35–37^. Laser capture microdissection (LCM) followed by liquid chromatography tandem mass spectrometry (LC-MS/MS) analysis of cellular tumor regions highlighted significant regional variability and numerous differentially encoded proteins (FDR = 0.1, n=3 separate microdissection replicates) (**Fig 2b-c, Supplemental Fig 5-6, Supplementary Table 1-2**). Pathway analysis, using a previous defined set of 64 proteogenomically concordance signatures^33^, highlighted regional heterogeneity in proliferative (MYC-, p=0.0006), invasive (KRAS-, p=0.03) and hypoxic (p=0.0048) programs^33^. Importantly, the enrichment of these programs was dampened when the entire tissue section was profiled; underscoring the value of regional profiling of highly heterogenous tumors using expression-based techniques (MYC-Red vs Bulk, p =0.001; Hypoxia-Red vs Bulk, p =0.008) (**Fig 2d-e**). Interestingly, the two major HAVOC partitions from Patient IV of the main study cohort showed a narrower profile of differences in functional programs (e.g. hypoxia), highlighting the capacity of HAVOC to detect even subtle molecular differences across individual cases driven by the microenvironment (p=0.04, t-test) (**Supplementary Fig 6b**).

Given the role of MYC in cell cycle progression, we validated these differences in the case presented in Figure 2b using CNN-based estimates of Ki-67 staining. This confirmed dramatic spatial variation in the proliferative capacity of these adjacent regions (**Fig 2f**, p < 0.0001). We further explored the predictive potential of HAVOC for heterogeneously proliferating subclones in another independent cohort of glioblastomas assembled to contain cases that displayed regional variations in Ki-67 (n=5, **Supplementary Table 2**). In all these cases, HAVOC’s heterogeneity maps captured H&E-based DLFV signatures that aligned and correlated with differential rates of proliferation (t-test, p<0.0001, **Supplemental Fig 7**).

To explore potential correlations between proliferation programs and morphology, we next assessed if specific DLFs were being activated by discernible histomorphologic features within regions displaying distinct Ki-67 indices in a representative case (**Supplemental Fig 8**). Indeed, FAMs of regionally-enriched DLFs highlighted a transition in pattern with areas with high cellularity (DLF214; activation on tumor nuclei) to regions displaying fibrillar cytoarchitecture (DLF134; activation on cytoplasmic processes) in variably high to low Ki-67 positive regions, respectively (**Fig 2g**). Interestingly, in this validation cohort, HAVOC-partitions (e.g. Orange, O) often formed “subclusters” (e.g. O1, O2, O3), that while geographically separated, showed similar morphologic patterns (e.g. hypercellularity, infiltrative phenotype) and maintained stable cluster-specific Ki-67 proliferation indices (**Fig 2h**). These findings suggested that H&E-based DLFV signatures may generalize to multiple spatially-separated tumor sub-regions and could help construct maps of biovariation even across larger specimens spanning multiple histopathological slides.

## Mapping biovariation across centimeter scale distances and entire specimens with HAVOC

Given the ability of hisomic signatures to group geographically separated tissue regions with similar biology on individual slides (Fig 2h), we next assessed the generalizability of this concept to the organization of tumor heterogeneity within individual large tissue specimens spanning multiple slides. As an example, we highlight a HAVOC map generated on a large recurrent isocitrate dehydrogenase (IDH)-mutated 1p19q-codeleted anaplastic oligodendroglioma, CNS WHO grade 3, showing heterogeneous radiographic signal and measuring 4.4 cm in maximum dimensions (**Fig 3a**). To map the histomorphologic heterogeneity across this entire case, we first generated seven sperate HAVOC partitions for each of the 12 H&E sections spanning the entire resected specimen (5.4 x 4.1 x 1.8 cm) **(Fig 3b)**. All 84 HAVOC-defined tumor regions were then histomorphologically organized across the entire specimen based on their pairwise DLFV similarities (*r*) (**Fig 3c**). Even across multiple sections, regions clustering more closely aligned with expert annotations rather than positional coordinates (**Fig 3d, Supplemental Fig 9**). This arrangement was further objectively validated with substantially different estimated proliferation indices (Ki-67) across the clusters of the geographically distributed partitions (p<0.005; Mann-Whitney U Test, **Fig 3d-e**). t-SNE mapping of all 10,973 0.27 mm^2^ image patches^38^ from this lesion also supported this HAVOC arrangement with a gradient of morphological and biological patterns, transitioning from nodular (blue), hypercellular (orange), moderately cellular (purple and green) and non-tumor/low cellularity (red) tumor regions (**Supplemental Fig 10**).

**Figure 3.**
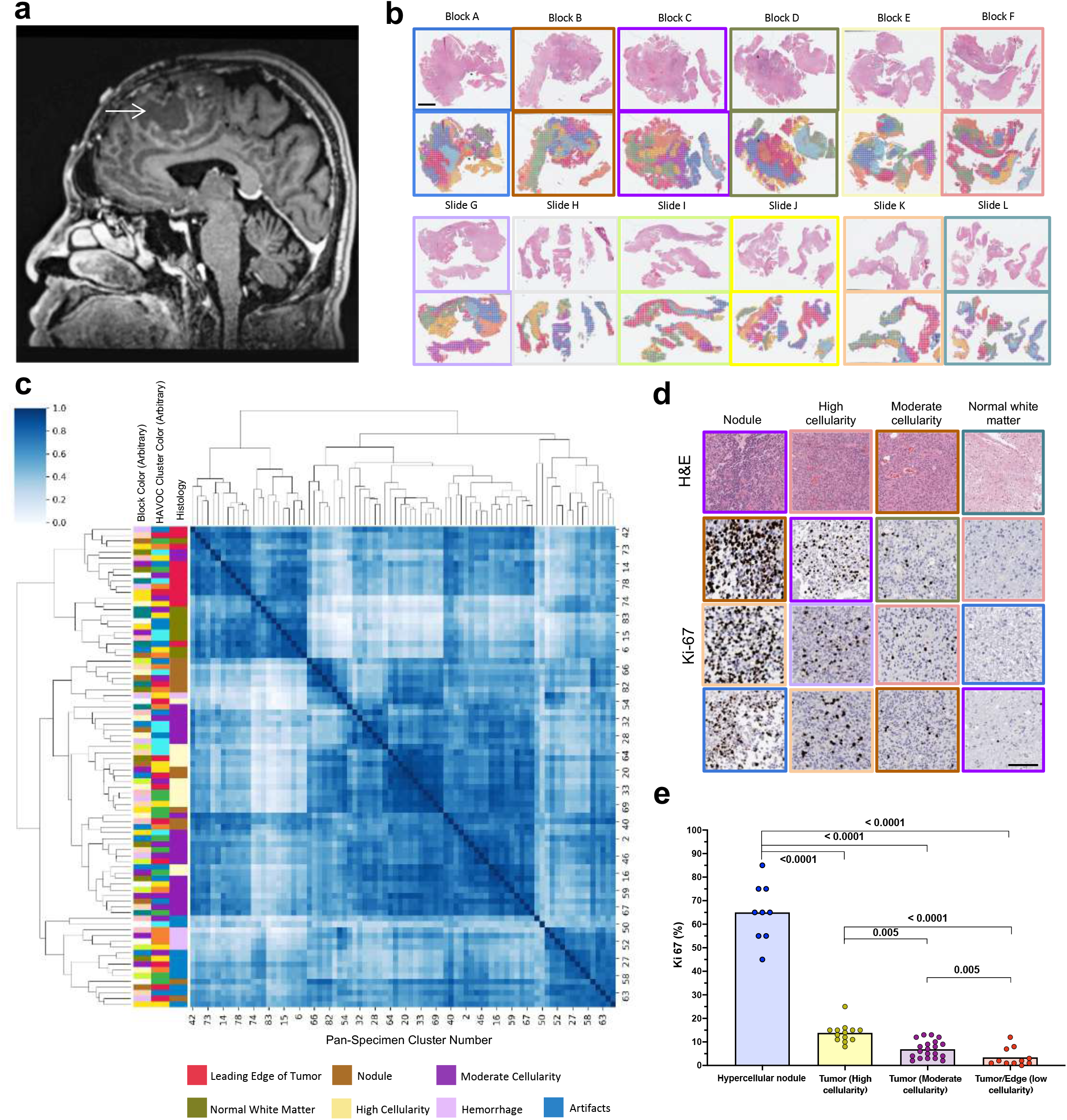
Small scale mapping of biovariation across entire tumor specimens using HAVOC. **(a)** MRI image of a recurrent IDH-mutated, 1p19q co-deleted oligodendroglioma measuring 5.4 x 4.1 x 1.8 cm in dimensions showing a heterogeneous pattern with multiple cysts and variable contrast enhancement. **(b)** 12 sequential H&E sections of the entire tumor with accompanying HAVOC heterogeneity maps. **(c)** Pairwise Pearson correlation matrix arranging all 84 clusters shown in panel (b). Slide colours corresponds to the same colours shown in former panel. There is a strong association of the clusters with expert-annotated morphologic patterns. **(d)** Representative images of Ki-67 (MIB1) from the different clustered regions. **(e)** Histogram of estimated Ki-67 (MIB1) across different regions.

To further examine if HAVOC could generalize to identify regional molecular profiles across larger tumor distances, we carried out this multi-slide mapping workflow on paired slide from three additional IDH-wildtype high grade gliomas (Histology: 2 WHO grade 4, 1 WHO grade 3) (**Fig 4a, d, Supplemental Fig 11, Supplementary Table 1**). Overall regional DLFV correlation mappings showed similar geographic relationships to global proteomic signatures (**Fig 4b,e**). In one case, HAVOC appropriately defined a focal hyperdense outlier region from other tumor morphologies (**Fig 4a-c**). In one of the other paired sets of samples, HAVOC partitions were reciprocally aligned with regions on different slides (**Fig 4d-e**). The final case also highlighted distinct diffuse infiltrating biology exclusive to only one of the two slides (Patient Va_RED), which was again appropriately segmented by HAVOC (**Supplemental Fig 11**). Interestingly, while quantification of regional cellularity provided further support for the proposed HAVOC groupings of the first 2 cases (**Figure 4)**, the third case pairing had a more uniform distribution of cellularity further supporting that HAVOC uses a diversity of cytoarchitectural features to guide clustering (**Supplementary Fig 12)**. All together, these experiments highlighted the potential for HAVOC to appropriately map and align intra-tumor areas of heterogeneity across large tissue distances.

**Figure 4.**
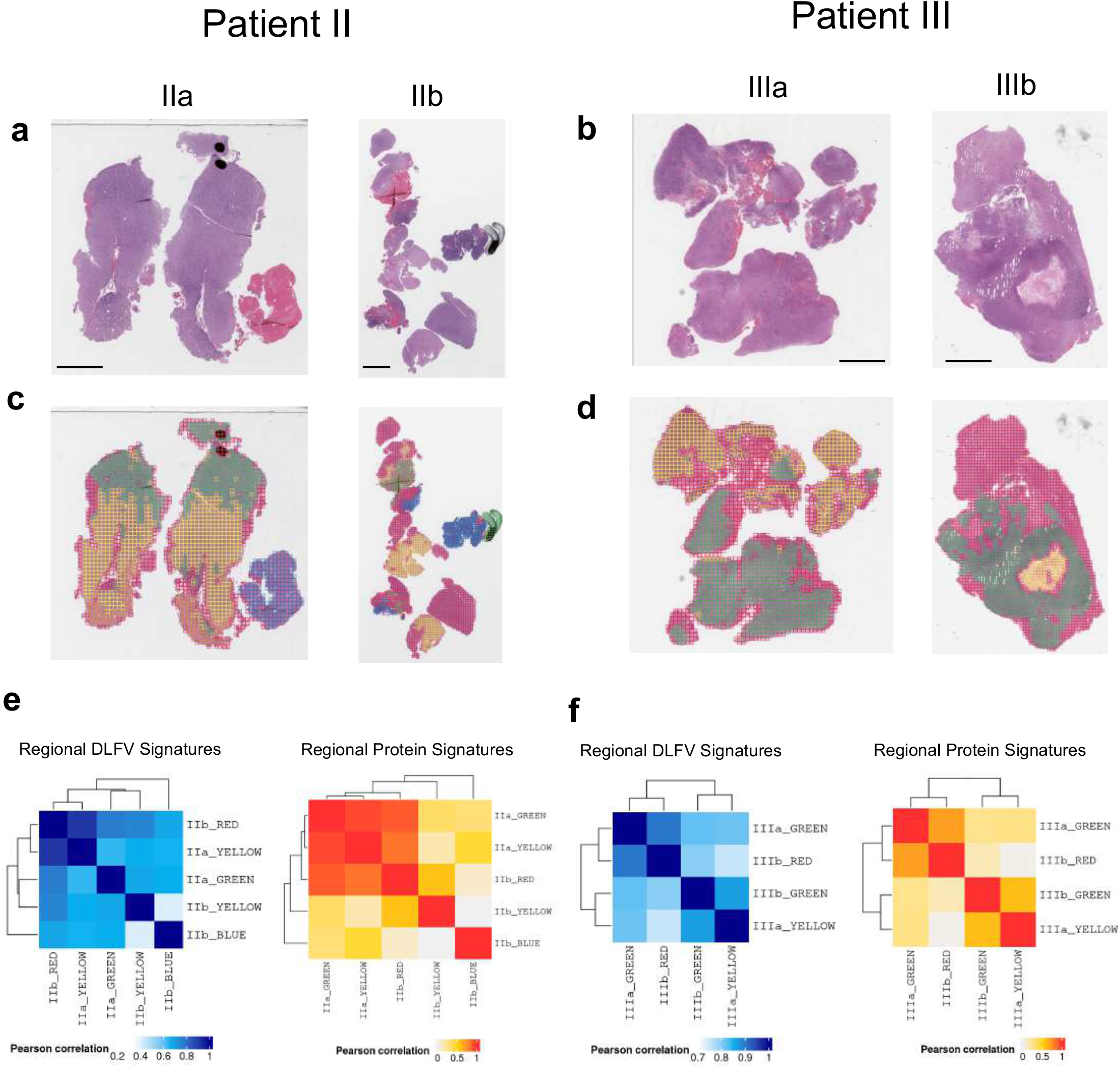
HAVOC mapping across independent slides align with overall molecular correlations. (**a**) HAVOC partitions across paired slides from an individual IDH-wildtype high grade glioma (**b**) Integrated DLFV correlation matrices across both slides define a distinct focal hyperdense outlier region (Sample IIb blue cluster). These multi-slide maps are in agreement with global patterns of proteomic variations derived from each of the major HAVOC defined partitions. (**c**) Another example of HAVOC multi-slide regional partitions that identified reciprocally aligned regions across slide pairs. (**d**) HAVOC-defined inter-slide niche similarities and agreement with proteomic variations.

## HAVOC reveals distinct geospatially separated differentiation states in high grade gliomas

Molecular descriptions of tumor biology are often defined based on sampling and profiling of a single tumor region^39^. We therefore next wanted to examine if HAVOC was detecting critical cellular states that may show reoccurring patterns of regional variation within individual tumors. We therefore aggregated the profiles of the 19 regions spanning six IDH-wildtype high grade gliomas (5 WHO Grade 4, 1 WHO grade 3; concurrently profiled in this study) and carried out a group analysis (**Supplementary Table 1**). Using the previously defined set of 64 proteogenomically concordant molecular programs, unsupervised clustering of the 19 regions was partly driven by patient ID; speaking to the high inter-tumoral heterogeneity found in IDH-wildtype high grade gliomas (**Supplementary Fig 13**). Importantly, this analysis also revealed spatial intra-tumoral variations in molecular programs in almost all profiled cases; including regional patterns of immune response, hypoxic response, proliferation and embryonic differentiation states. UMAP dimensionality reduction revealed a significant influence of the later differentiation signatures across the entire cohort (e.g. genes associated with an embryonic “poorly” differentiated cell state^40^) (**Fig 5a**). This was further supported with a strong inverse correlation with regional programs associated with a mature astrocytic phenotype (r=0.71, p <2e^-16^) (**Fig 5b**). Supervised regional analysis of this astrocytic-embryonic differentiated axis found that at least 5 out of the 6 profiled cases showed significant spatial differences (p <0.005, ANOVA) (**Fig 5c**). We found this high co-occurrence of these contrasting patterns notable, as tumors, across multiple organ sites, are often still approximated as being either “poorly” or “well” differentiated upon clinical presentation.

**Figure 5.**
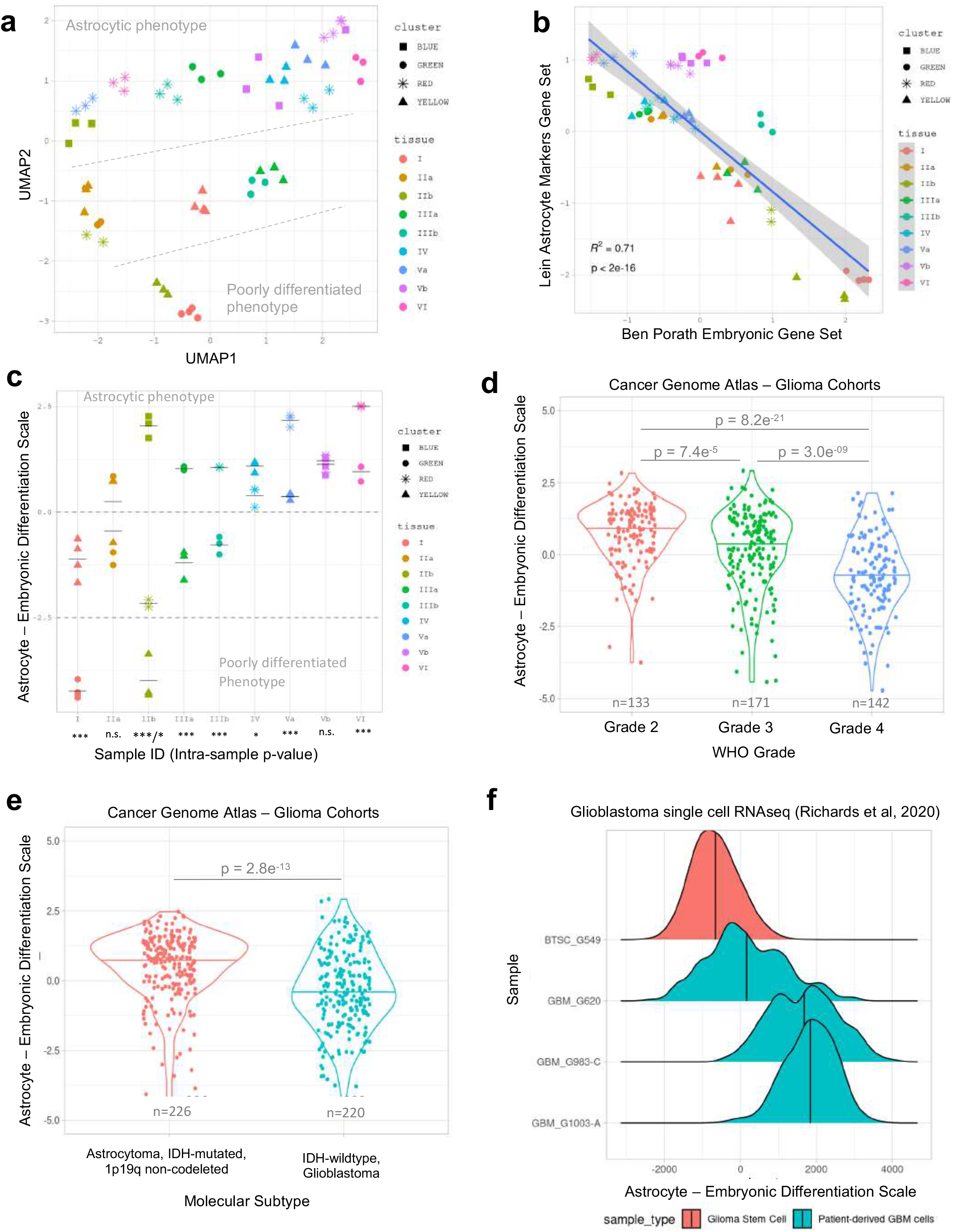
HAVOC reveals spatially organized patterns of molecular heterogeneity in high grade gliomas. (**a**) Unsupervised analysis of 19 HAVOC defined tumor regions across 6 high grade gliomas by performing UMAP dimensional reduction of the ssGSEA scores of 64 proteogenomically concordant genesets (**b**) Inverse relationship between the ssGSEA scores of the embryonic stem cell state (Ben Porath ES_1) and the Astrocytic differentiation geneset (Lein Astrocyte Markers Gene Set). p-value generated by linear fit model. (**c**) Regional differences in state of differentiation (Astro-ES axis) across each of the spatially resolved and profiled slides. The level of statistical significance of the differences between the regions of each tissue slide was assessed by ANOVA; p-values are indicated as follows: *** < 0.001, ** < 0.01, * < 0.05, and n.s. = not significant. (**d**) Distribution of the Astro-ES axis in astrocytic tumors (non-1p19q codeleted) from TCGA-GBM and TCGA-LGG cohorts stratified by WHO grade and (**e**) IDH status. (**f**) Varying distributions along the Astro-ES axis at the single-cell level in patient-derived GBM cells (n=3) and glioma stem cells (n=1); dataset previously published by Richards et al.

In the Cancer Genome Atlas (TCGA), this differentiation axis correlated with aggressiveness with both lower WHO grades (p < 7.4e^-5^, n=446) and IDH mutations (p = 2.8e^-11^) of astrocytic (non 1p19q-codeleted) tumors showing significantly higher differentiation signal at the bulk level (**Fig 5d,e**). We also found support for both inter- and intra-tumoral variation along the astrocytic-to-embryonic axis in single cell RNA data generated from both patient-derived glioblastoma tissue specimens and brain tumor stem cell cultures (BTSCs)^41^ (**Fig 5f, Supplemental Fig 14**). Interestingly, while some samples analyzed showed complex expression patterns with multiple co-existing states of differentiation (GBM_G620; GBM_G983-C), others displayed fairly homogenous signatures (GBM_G1003-A); presumably related to both the sampled region and the overall tumor biology. The notable spectrum of steady-state positions in differentiation states in expanded BTSCs suggests that generation of patient models from single individual tumor regions may only partially capture potential region-to-region variations in differentiation uncovered in this analysis. Taken together, these results highlight how neural network-guided multi-regional sampling and profiling of cancer tissue can reveal new insights of biovariation that may be hidden and not immediately accessible from alternative single region profiling strategies.

## HAVOC biodiversity maps generalize to genomic differences and untrained tissue types

As tumor heterogeneity is a challenge relevant to many cancer types^42^, we next assessed the generalizability of HAVOC to other non-central nervous neoplasms. First, we retrieved an available WSI of an experimental metastatic lung cancer mouse model in which two independent clones of a lung tumor were injected into the tail vein and subsequently resulted in multiple spatially distinct liver metastases^6^ (**Fig 6a**). Importantly, using spatial DNA and RNA sequencing, two of the five metastatic deposits, in addition the intervening liver tissue, were characterized in the originating study using copy number and expression patterns differences (**Fig 6b, Supplemental Fig 15a**). Using HAVOC, we generated 11 total partitions (to ensure saturation of the different histomorphological patterns) and compared them with the characterized ground truth annotations. Indeed, image-based clustering segmented all five tumors into fairly homogeneous lesions, in addition to defining various regional histological patterns of the liver tissue (**Fig 6c-d, Supplemental Fig 15b-c**). Using the silhouette method, these five tumors formed two major clusters (k=2 subclones) as the most parsimonious solution (**Supplemental Fig 15d**). Notably HAVOC separated out both the two genomically-distinct subclones and peritumoral liver tissue with and without inflammation, further supporting that distinct histomorphologic fingerprints can be leveraged to predict regions of potential genomic, transcriptomic and cell composition heterogeneity (**Fig 6d**). Subsequent FAMs exploring morphologic correlates highlighted distinct histomorphologic patterns within the tumor subclones defined by the original study. In Clone A, FAMs highlighted a fairly advanced organization of tumor cells with abundant nuclear palisades, while Clone B showed a more random arrangement with large, atypical cells scattered throughout the lesion. Further, FAMs of the peritumoral liver tissue highlighted a differential distribution of inflammatory cells across HAVOC-proposed groupings, in agreement with the tumor, normal, and immune cell classes assigned from the original single-cell slide-RNA-seq projections (**Fig 6e**).

**Figure 6.**
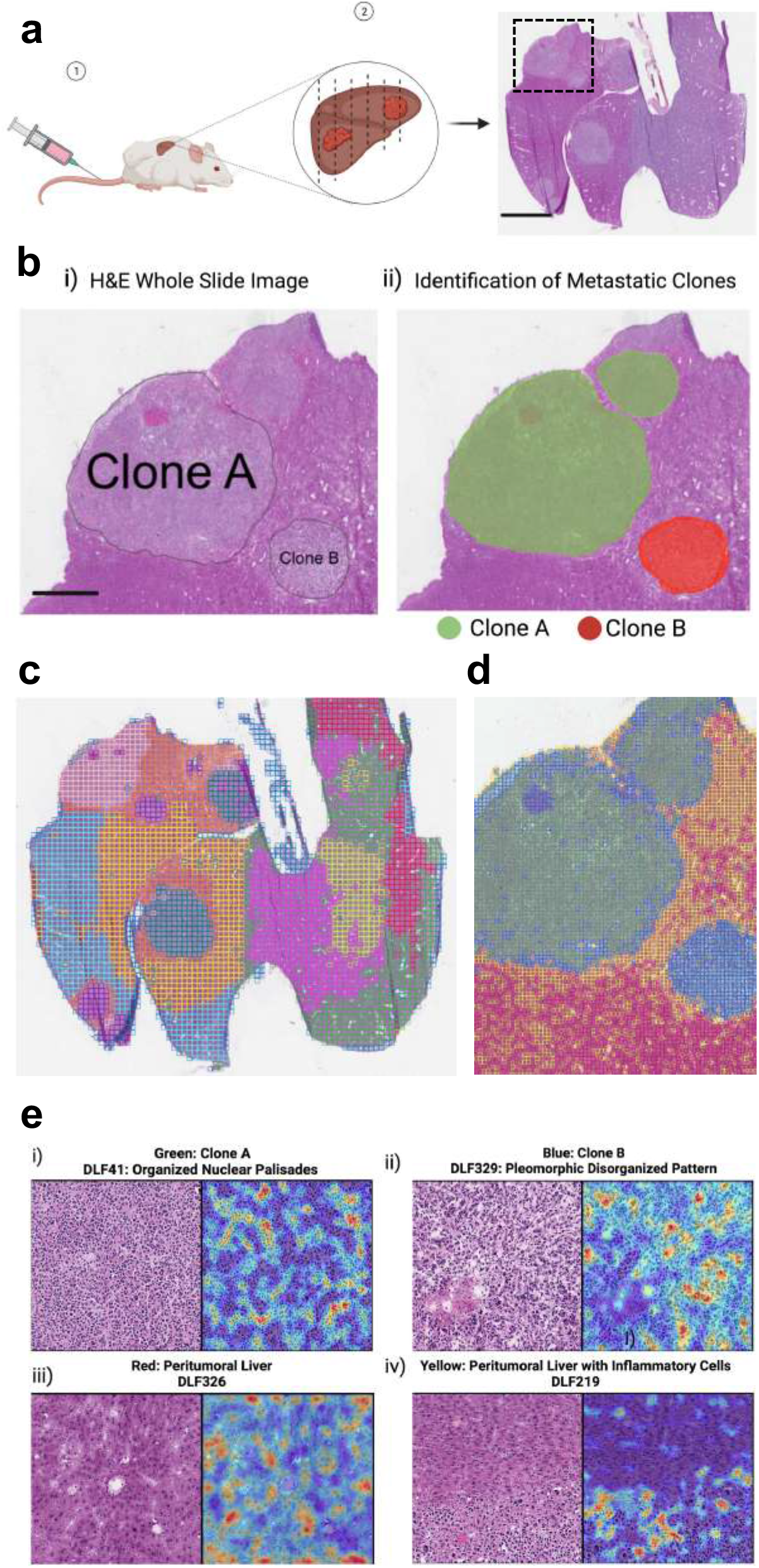
HAVOC defines genetically distinct metastatic clones. **(a)** Schematic and H&E-stained section of a mouse liver modeling polyclonal Kras^G12D/+^Trp53^*-/-*^ lung cancer metastases from *Zhao et al*. Spatial profiling in the boxed region allowed benchmarking of HAVOC in this model. Scale bar: 5 mm. **(b)** Slide-DNA-seq and Slide-RNAseq of focused region provided ground truth of normal liver, tumor clone *“*A*”* and *“*B*”*. Dotted lines in the right panel indicate tumour boundaries. Scale bar: 2 mm. **(c)** HAVOC mapping of entire H&E sections with 11 partitions to ensure saturation of the different histomorphological patterns. HAVOC segmented all five tumors into stable groupings, identified in pink, teal, and purple. **(d)** HAVOC partitions (k=4; tile width: 128 pixel) of the focused region mapped by slide-DNA seq (original study) spatially divided tumor into 2 homogeneous subclones (green vs blue). Surrounding liver tissue was also separated into regions of peritumoral liver and immune infiltrates (red vs yellow) that match single-cell RNA seq projection (see supplemental Fig 13a for ground truth from original study). **(e)** FAMs of selected differentially activated DLFs for each tumor regions (i-ii) and DLFs enriched in peritumoral region highlighting liver regions with and without inflammatory cells (iii-iv).

While the other three remaining tumor regions were not genomically annotated in the original study (potentially due to cell throughput limitations of the spatial DNA sequencing technique), they did indeed show similar histologic patterns to the more atypical tumor, confirming the ability of HAVOC to group metastases of similar histomorphologies, even across entire organs.

To further test the generalizability of the HAVOC workflow to other untrained tissue types, we next applied it to “interesting” dermatopathology and pulmonary pathology cases showing a squamous neuroendocrine “collision” tumor and divergent adenosquamous tumor differentiation respectively (**Fig 7**). On the skin biopsy, the position of the squamous cell carcinoma (*in situ)* is highlighted by the high molecular weight keratin (CK34BE12). Within the dermis, a distinct infiltrative neuroendocrine neoplasm composed of sheets, nests and cords of round basaloid cells that labeled with synaptophysin. Indeed, HAVOC-proposed groupings of this specimen distinguished the distinct squamous and neuroendocrine morphologies in alignment with the distinct immunohistochemical staining (**Fig 7a-b**). HAVOC partitioning of the case of adenosquamous carcinoma also showed a high spatial concordance to TTF1 (adenocarinoma) and p40 (squamous carcinoma) immunopositive tumoral components (**Fig 7c-d, Supplementary Fig 16**). Altogether, these data demonstrate that HAVOC can serve as a tissue type- and molecular-agnostic tool to map biodiversity in different cancers.

**Figure 7.**
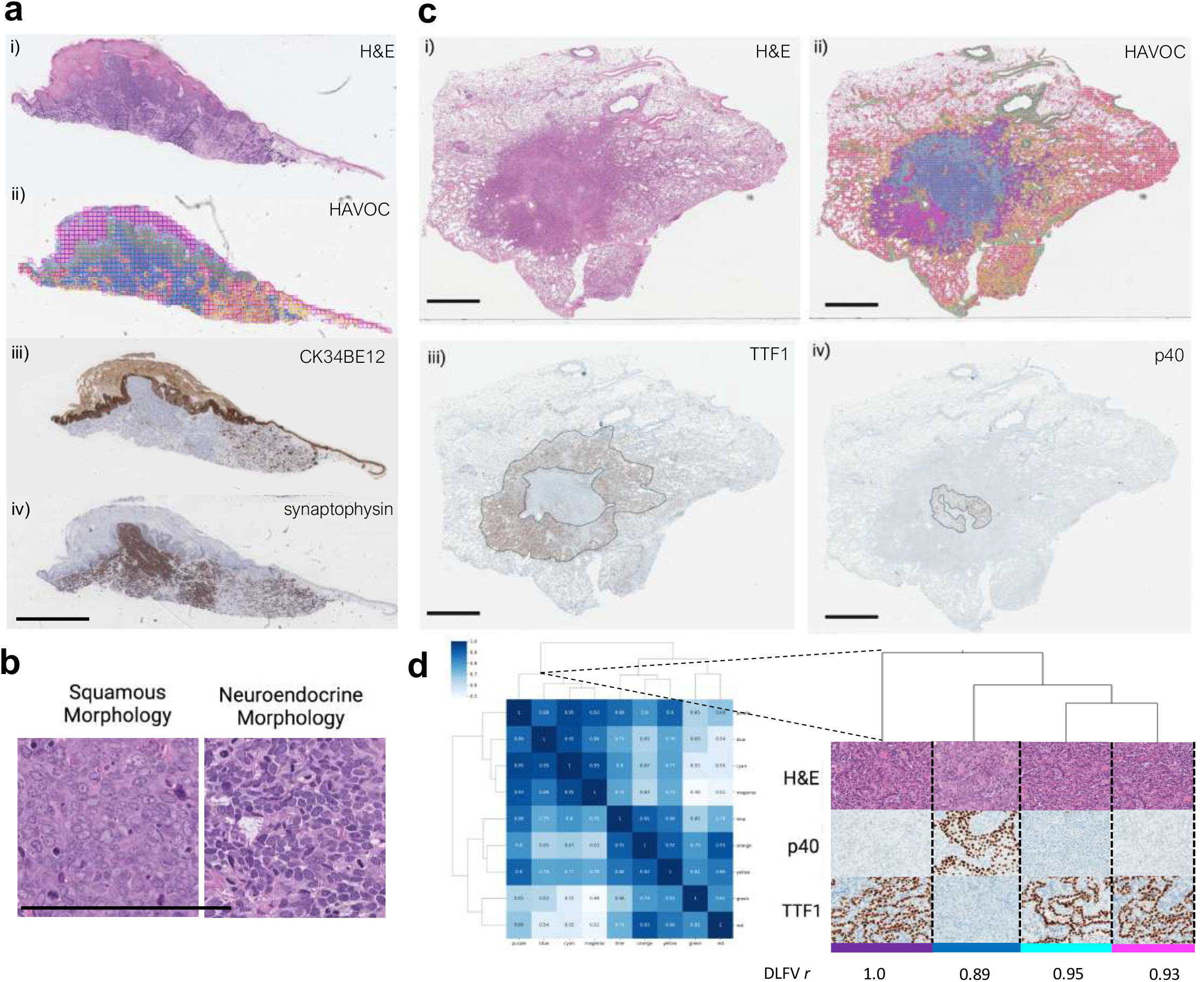
HAVOC generalizes to untrained tissue types. **(a)** (i) Dermatopathological specimen of a “collision” tumor comprised of distinct regions of squamous cell and neuroendocrine carcinoma. (ii) HAVOC partitions resolved the distinct tumor types that matched the immunohistochemical ground truth (iii) CK34BE12 and (iv) synaptophysin. Scale bar: 2 mm. **(b)** Representative high power magnification micrographs of HAVOC-defined distinct tumoral sub-clones in dermatopathological specimen highlighting the squamous (green partition) and neuroendocrine (blue partition) components. Scale bar: 200 *μ*m. **(c)** (i) H&E-stained lung resection with a neoplasm showing divergent adenosquamous differentiation. Tumor sub-components are highlighted with distinct (ii) p40 (squamous) and (iii) TTF1 (adenocarcinoma) immunohistochemical staining. (iv) HAVOC partitions of original WSI show subregions that align with the distinct tumoral patterns defined by the squamous/adenocarcinoma markers. Scale bar: 6 mm. **(d)** Hierarchical clustering of HAVOC-defined regions and representative (i) H&E, (ii) p40, and (iii) TTF1 images reveals two distinct tumor sub-patterns.

## Discussion

Intra-tumoral heterogeneity has emerged as a key concept in current precision medicine efforts^42^. While there have been technological breakthroughs to map such spatial biodiversity at the genomic, transcriptomic and proteomic level, the relative discord between the upper limits of tissue profiling throughput and tumor sizes creates an important bottleneck for comprehensive analysis of most tumor specimens^2,4,7^. Here, we highlight how a neural network-based tool (HAVOC) can detect and quantify spatial distributions of cancer biodiversity across not only individual sections, but also on larger (entire) tumor specimens in a hypothesis-agnostic manner. Importantly, we go to great lengths to prospectively validate that these proposed histomorphologic patterns of spatial heterogeneity correlate with expert human interpretations, key biomarkers of aggressiveness and global genetic, transcriptomic and protein-level differences. Using this tool, we partitioned and profiled 19 distinct regions from 6 IDH-wildtype high grade gliomas specimens and uncovered that different spatially confined states of differentiation can co-exist within the same tumor. Interestingly, while the defined poorly differentiated state correlated with many important indicators of clinical aggressiveness (WHO grade and IDH status), it does not appear to correlate with proliferation or tumor initiating potential in previous studies, adding another layer of complexity to existing models of tumor heterogeneity. Importantly, such findings also underscore the potential of histomic mapping to guide and improve representative sampling and characterization of heterogeneity across entire tumor resection specimens. While we focused our discussion and validation efforts on spatial patterns of differentiation, there were many additional case-by-case geographic differences identified that were outside the scope of this study (e.g. proliferation, hypoxia, immune response). We therefore make this unique morphology-driven spatial analysis of the glioma proteome available for further exploration through an inter-active data portal (Brain Protein Atlas^29^; https://www.brainproteinatlas.org/dash/apps/ad).

There are important distinctions of how HAVOC complements other computational approaches to mapping intra-tumoral molecular heterogeneity directly from histology. Approaches that aim at directly training and/or correlating deep learning outputs to specific molecular events are inherently empirical and may be most effective at predicting fairly robust molecular signatures that are driven by heterogenous cellular compositions (e.g. presence of lymphocytes, vessels) and/or a handful of transcriptional profiles (e.g. cell division)^22,23^. While these may be highly effective and cost-efficient in some clinical and research contexts, such approaches may not have capacity to extend and generalize across all cancer types and relevant molecular pathways of interest. This presents an important limitation and gap for targetable biological programs that may not have strong “histomolecular” correlates, cancers with a high degree of heterogeneity and for more modern personalized medicine strategies^27,28^. A more hypothesis-free approach has been the use of supervised deep learning approaches to identify specific patterns within tumors (e.g. areas devoid of tumor infiltrating lymphocytes and stromal elements) that may signal differential biology^14^. These “at risk” regions provide spatial targets for prospective and personalized profiling of individual cancer specimens. We believe HAVOC extends this concept further by decoupling the need for any pre-defined feature that may potentially bias or limit exploration into mapping of biovariation with associated phenotypic changes. While, in certain circumstances, this may compromise the sensitivity for detecting subtle spatial variations in important molecular programs, we believe it provides a highly dynamic and generalizable solution for personalized discovery and characterization of spatial changes in tumor biology. It therefore can be directly extended to not only a variety of tumor types, but even to the longitudinal analysis of cancer in which molecular patterns may be influence by both tumor progression and treatment-related effects^9^.

HAVOC does not require any dedicated tissue or significant computational resources, and contains only a handful of tunable parameters (e.g. tile size, cluster numbers) that can be easily adjusted depending on the specific context (e.g. smaller tumor deposits). These capabilities, when combined with the processing of serial sections for molecular analysis, provide a computational approach for the characterization of intra-tumoural heterogeneity at practically any scale and level of resource availability. Moreover, as we demonstrated in this study, the large diversity of images used to train the default CNN within HAVOC provides an adaptable unsupervised framework to detect tumor heterogeneity in a relatively species-, tissue-, molecular platform-agnostic manner. We, therefore, envision HAVOC to serve as a routine and powerful research tool for mapping intra-tumoral heterogeneity across both large-scale and more intricate (n-of-1) tissue profiling studies. Given its ease of implementation and compatibility with FFPE sections, we envision HAVOC becoming an essential tool to map heterogeneity on all clinical and research tissue-based to help provide more systematic approaches to tissue selection for ongoing personalized precision medicine efforts.

Because HAVOC is built to detect histomorphologic pattern differences, it can be theoretically extended to non-neoplastic and immunohistochemistry-resolved tissue heterogeneity that may be niche- and epigenetically patterned and independent of genetic (e.g. copy number) alternations. Moreover, when coupled with salient feature activation mapping, it can complement spatial transcriptomic and proteomic approaches by providing phenotypic correlates (e.g. edema, increased nuclear: cytoplasmic ratio) to explain/predict interesting expression-level differences. To facilitate wide adoption, we have packaged HAVOC in a number of ways to promote ease of use. For large-scale initiatives, we provide source code to allow HAVOC to be deployed locally on large cohorts in an automated manner. For more translational researcher and clinicians, we also host HAVOC in a cloud-based server (https://www.codido.co) that allows analysis of digitized slides (.SVS) without any need for advanced software expertise or hardware. There are some important caveats of HAVOC that should be considered when using this tool. Firstly, its fundamental dependence on tissue patterns means that it can be influenced by non-biologically factors (e.g. tissue folds/tears/focus/suboptimal uneven staining). In our experience, we found that these artifacts can be easily identified and excluded from downstream analysis; both by careful post-hoc analysis or by supervised classification of clusters to label tumor-specific partitions. Moreover, the sensitivity of HAVOC for very subtle and intermixed patterns of biodiversity likely also needs to be assessed in a context-specific manner. These biological factors may dictate the particular type of molecular profiling technique best suited for HAVOC pairing. In summary, HAVOC provides a highly flexible, generalizable, accessible, and scalable approach for mapping histomorphologic-correlated phenotypes and could serve as an essential tool to explore and document biologic heterogeneity in human tissue and disease.

## Data Availability

The mass spectrometry proteomics data of all HAVOC derived regions presented in this manuscript have been deposited to the ProteomeXchange Consortium via the PRIDE partner repository with the dataset identifier PXD037548 (username; password: fy4uPFAi). The data can also be examined directly through our interactive data portal (Brain Protein Atlas29; https://www.brainproteinatlas.org/dash/apps/ad). Labeling legend for mass spectrometry proteomics data can be found at https://bitbucket.org/diamandislabii/havoc. As described, some of the data used in this publication derived from The Cancer Genome Atlas Program (TCGA) and deposited at the Data Coordinating Center (DCC) for public access [http://cancergenome.nih.gov/]. The RNA-Seq IvyGAP data used are publicly available at Gene Expression Omnibus through GEO series accession number GSE107560. The single-cell are publicly available through the Broad Institute Single-Cell Portal (https://singlecell.broadinstitute.org/single_cell/study/SCP503) and CReSCENT60 (https://crescent.cloud; study ID CRES-P23). Additional proteomic data from different glioblastoma regions were also derived from Lam et al are also publicly available through the ProteomeXchange Consortium via the PRIDE partner repository with the dataset identifier PXD019381. The H&E slide and ground truth annotations for the metastatic lung carcinoma mouse model was provided directly from the authors and the relevant study.

## Author contributions

A.D., K.F., S.L, A.O and P.D. conceived the idea and approach. K.F. developed the computational workflow. A.D. and B.L. designed and carried out molecular validation experiments. N.A., P.P. A.G., Z.S.K., P.D. provided relevant cases and pathological annotations for the various validation studies. A.L. and Q.T. performed bioinformatic analysis. A.D. K.F. and P.D. wrote the manuscript, with input from all other authors.

## Acknowledgments

The Diamandis Lab is supported by the Terry Fox New Investigator Award program, the Canadian Institute of Health Research and the Brain Tumor Foundation of Canada. A.O. S.L., P.D, and P.P also received research grant support from the Princess Margaret Cancer Foundation and the Ontario Institute for Cancer Research.

## Competing Interests

The authors declare no competing interests.

## Methods

### Ethics statement

The University Health Network Research Ethics Board (UHN REB) approved the study REB #17-6193 as it has been found to comply with relevant research ethics guidelines, as well as the Ontario Personal Health Information Protection Act (PHIPA), 2004. Patient consent was not directly obtained and instead a consent waiver for this study was granted by UHN REB as the research was deemed to involve no more than minimal risk as it used exclusively archival tissue specimens.

### Tissue cohort development and digital scanning

All clinical cases included in our study cohort were retrieved from UHN archival tissue specimens. To assess potential associations between DLFV correlation coefficients and human-perceived differences, an initial brain tumor tissue cohort (n=40) largely comprising of diffuse glioma cases (IDH-wildtype and IDH-mutants, WHO Grade 2-4) was developed. To compare proteome-level programs between distinct partitions, cases were further examined to define regions relatively pure in tumor content (e.g. not regions of low tumor purity) and with partitions sufficiently large enough for laser capture microdissection (LCM) and LC-MS/MS analysis. In total, the LC-MS/MS cohort included seven high-grade gliomas^31^. Six cases had glioblastoma histology and an IDH-wildtype immunohistochemical staining pattern. The final case was an IDH-wildtype high grade glioma with a WHO grade 3 histology. Age ranges were provided for anonymity and patient IDs were not known to anyone outside of research group (**Supplementary Table 1**). Additional independent confirmatory cases were also selected which displayed region-to-region heterogeneity in their MIB1/Ki-67 proliferation indices (n=5 independent cases). The collision tumor in the skin and the lung adenosquamous carcinoma were also retrieved from our local cancer center (UHN). The H&E section of the metastatic lung cancer model was provided directly from the authors of the relevant study^6^. The entire generated local cohort was digitally scanned as WSI with a compression quality of 0.70 and a magnification of x20 on a Leica Aperio AT2 scanner.

### Implementation of the deep convolutional neural network

We used a previously trained pathology-optimized version^32^ of the VGG19 CNN^43^ to extract the deep learning feature vectors used during HAVOC partitioning^30^. Specifically, the original VGG19 model was optimized using transfer learning on a set of 838,644 pathologist-annotated image patches spanning over 70 brain tissue and tumor types derived from over 1000 patients^32^. We previously showed that this training diversity lead to node-weights within the network that align with human-discernible histopathological feature representations that are applicable across multiple malignancies^15,32^. Given the unsupervised nature of HAVOC, the CNN was used without having to undergo any additional training or optimization for this application. Previously learned class labels from the original fine-tuning of the VGG19 were decoupled from this analysis and instead only the feature representations, in the form of a 512-dimentional vector from the global average pooling layer of the CNN, prior to SoftMax reduction and classification, were extracted and used for image-based clustering^32^.

### Regional partitioning and estimation of heterogeneity

Tissue partitions by HAVOC were generated using an unsupervised image clustering framework as previously described^15^. Briefly, WSIs for each clinical specimen of interest are first tiled into individual 0.066-0.27 mm^2^ image patches. The particular size of the image patched was largely dictated by the size of the available tissue (smaller tiles for smaller tissue) and histomorphologic pattern of interest (e.g. cytoarchitecture vs. nuclear patterns benefiting from larger (low power) vs. smaller (high power magnification) tiles respectively). Histomorphologic fingerprints for each tile are then generated by averaging the Deep Learning Feature (DLF) values from the final global average pooling layer of the VGG19 CNN. We refer to this 512-dimentional feature representation matrix as the Deep Learning Feature Vector (DLFV). As this network was tuned on a diverse set of histological images, many individual DLFs align with human-identifiable histologic features, including fibrosis, epithelium, and mucoid patterns thus allowing it to group images patches, with relatively similar morphologies, into meaningful clusters^32^. To identify spatial transitions in histomorphological patterns for each case, the DLFVs generated from each tile are clustered, using Ward hierarchical clustering, as previously described^32^. Images that group together by specific clustering threshold (default k=9) are deemed to show a relatively similar histomorphologic signature and grouped together to form a HAVOC partition. In our experience, these often tend to show spatial relationships concordant with expert histopathologic review. The relative distances (similarity) of each HAVOC partition (spatial region) are quantified using the Pearson correlation coefficient of each region’s average DLFVs. In addition to quantitatively correlating with histomorphologic differences perceived by humans, we found that the overall correlation coefficient is less affected by artifacts that often skew results when raw DLFV were used to cluster regions spanning multiple slides. Therefore, while we find clustering of raw tile DLFVs from individual slides effective, we observe more robust results with correlation coefficients are used to compare HAVOC partitions from independent slides.

### Region-specific deep learning feature selection and mapping

To understand which morphologic features were potentially driving, or at the very least associated with, the unsupervised slide subgroupings, we first identified the most significantly enriched DLFs in each of the HAVOC-proposed groupings. To visually determine what morphology such DLFs were detecting, we then assess image tiles at the extremes of the DLFs of interest across the entire slide (both the highest-scoring and the lowest scoring). Interpretations of potential morphological differences between the tiles at the two extremes were provided by two or more blinded pathologists. This qualitative expert assessment was then complimented and validated by generating isolated feature activation maps (FAMs) of the candidate DLFs of interest to evaluate if the activated tile coordinates matched the location of the histologic feature suggested by expert pathologists. Python code used to generate the FAMs can be found at BitBucket (https://bitbucket.org/diamandislabii/faust-feature-vectors-2019).

### Pathologist evaluation of HAVOC-defined heterogeneity

Both computational silhouette and human evaluation approaches were employed to evaluate the pathologic correlations of HAVOC-proposed regions of heterogeneity and determine the optimal number of WSI partitions when appropriate. For most cases, we found that the vast degree of pathologist-discernible patterns of histomorphologic variations reached saturation after 7-8 WSI partitions irrespective of the overall level of complexity and tissue heterogeneity. While 7 partitions worked well for mapping most histomorphologic patterns of variation in our clinical cohort, the clustering framework is easily tunable and can be extended to addition partitions using a stepwise k-means clustering approach to explore additional and more subtle regions of heterogeneity in complex tissue specimens (**Supplemental Fig 1, 15 & 16**). To capture finer microscopic details, relevant in some circumstances, the size of the image patches can also be tuned to the appropriate application. For examination of relatively large regions of cellular tumor, image patches with a width of 512 and 1024 pixels works extremely well (**Fig 2**). For more focal deposits (**Fig 6**), the tile size can be reduced to 256 or 512 pixels in length to reduce the intra-patch pattern variation.

### Laser capture microdissection (LCM) and LC-MS/MS proteomic profiling

Tissue sections were stained prior to LCM to improve contrast and provide references across the slides to guide precise microdissection as previous described^33,44^. Specifically, formalin-fixed paraffin-embedded (FFPE) tissue blocks were sectioned (at 10 um thickness) and mounted onto Leica PEN slides (Cat No. 11505189). Slides were subsequently deparaffinized with 100% xylene (2x), 100% ethanol, 95% ethanol, 70% ethanol, and 50% ethanol (3 minutes each). These slides were then stained with hematoxylin (1 minute), rinsed in de-ionized water (1 minute), and stained in 1% eosin Y (Fisher scientific). Slides were finally air dried prior to laser capture microdissection.

Regions of interest from these sections were micro-dissected using a Leica LMD 70000 (Leica Microsystems, Inc., Bannockburn, IL). HAVOC-generated color tiled maps were used as a reference to guide parameters for dissection. Samples were collected in an Eppendorf tube and subsequently stored at room temperature until further sample preparation began.

Proteomic extraction was performed with the addition of 50 uL of 1% Rapigest, 200 uL of a dithiothreitol, ammonium bicarbonate (50 mM), and tris-HCl solution to each sample. Samples were subsequently sonicated using a Bioruptor Plus on high with 30 second intervals for 1 hour. Solutions were heated to 95 degrees Celcius for 45 minutes, followed by 80 degrees Celsius for 90 minutes with a ThermoMixer. 20 uL of iodoacetamide was then added to each solution in the absence of light for alkylation. 1 ug of trypsin/Lys-C mix was then added to each sample and reacted overnight at 37 degrees Celsius. The solutions were subsequently acidified with trifuoroacetic acid at a final concentration of 1% ahead of stagetip cleanup.

In preparation for mass spectrometry analysis, samples were desalted using Omix C18 tips following manufacturer protocol. Elution of peptides was completed with 3 uL (0.1% formic acid, 65% acetonitrile) and dilution of peptides was completed with 57 uL (0.1% formic acid in MS water). 18 uL of solution (2.5 ug of peptides) was loaded with an autosampler for mass spectrometry as previously described^33^. Briefly, peptide elutions from an EASY-Spray column ES803A occurred at a rate of 300 nL/min with increasing concentration of 0.1% formic acid in acetonitrile over a one hour gradient. This setup was coupled to a Q Exactive HF-X with a spray voltage of 2 kV with a 60 minute data-dependent acquisition method. The full MS1 scan was completed from 400-1500 m/z at a resolution of 70, 000 in profile mode, with the top 28 ions selected for further fragmentation with HCD cell. Fragment detection occurred with an Orbitrap using centroid mode set at a resolution of 17,500. The following MS parameters were used: MS1 Automatic Gain Control (AGC) target was set at 3 × 106 with maximum injection time of 100 ms, MS2 AGC set at 5 × 104 with maximum injection time of 50 ms, isolation window of 1.6 Da, underfill ratio 2%, intensity threshold 2 × 104, normalized collision energy (NCE) of 27, charge exclusion was set to fragment 2+, 3+ and 4+ charge state ions only, peptide match was set to preferred and dynamic exclusion set to 42 (for 90 min method).

### Entire tumor histomic profiling and organization

HAVOC evaluation of larger and even entire tumor specimens was completed by histomic profiling of each H&E-stained section for each of the corresponding tumor block. Unsupervised analysis of each WSI was completed individually as described in previous sections and then merged in a separate and final step. For the example of the 12-slide tumor specimen shown, the number of clusters for each WSI was set to 7; within the range that results in saturation of perceivable histomorphologic patterns at a tile size of 0.27 mm^2^. The DLFVs of each cluster from each of the related WSIs of the specimen were then subsequently organized using pairwise Pearson Correlation coefficients. We provide a separate script for this final step in the repository. Multi-slide Pearson correlation matrixes of HAVOC proposed groupings were generated using Matplotlib and Seaborn Python data visualization libraries. Evaluation of the fidelity of the multi-slide regional alignment was assessed by clustering of expert annotations of each HAVOC-partition, immunohistochemical staining patterns (MIB1/Ki-67) and LC-MS/MS analysis for the 3 patients with paired slides.

### Statistical Analysis

MaxQuant Andromeda (version 1.5.5.1) search engine was used to process mass spectrometry raw data files against the Human Swissprot protein database (July, 2019 version). Proteins were filtered to include only those appearing in at least 60% within a sample. Raw protein values were Log2 transformed, with non-valid values imputed (downshift=0.3, width=1.8). Analysis of proteomic data was performed using biostatistical platforms Perseus (www.coxdocs.org) and ssGSEA^45^ (https://gsea-msigdb.github.io/ssGSEA-gpmodule/v10/index.html). Single sample gene set enrichment analysis was used to define pathways enriched in each HAVOC proposed region. One-way ANOVA testing and Tukey’s post hoc test were completed in R (v4.0.4) to identify differences in enriched pathways across HAVOC proposed groupings. Student’s t-tests were used to test the difference in Ki-67 proliferation indices across HAVOC proposed groupings. Additional statistical tests used for individual analyses are mentioned in the appropriate text and figure coordinates.

### Gene set enrichment analysis

Gene sets enrichment analysis was used to understand the biological significance of the regionally distinct molecular profiles both on individual samples (**Fig 2**) and at a cohort level (**Fig 5**). Three gene expression datasets were analyzed: (i) Proteomics dataset generated in this study. For this, proteins with >25% of missing values were removed, resulting in a dataset of LFQ intensity values with 1,920 proteins across 60 injected aliquots, which typically included 3 technical replicates per tissue specimen. (ii) RNA-seq data and associated clinical information from the merged cohort of LGG and GBM (Ceccarelli et al, *Cell*, 2016) retrieved from cBioportal (http://www.cbioportal.org/study/summary?id=lgggbm_tcga_pub). Oligodendrogliomas were excluded from the analysis resulting in a cohort 446 diffuse “astrocytic” tumors. (iii) The final cohort represented normalized scRNA data previously published by Richards et al. (*Nature Cancer*, 2020). This dataset was downloaded from the Broad Institute’s Single Cell Portal (https://singlecell.broadinstitute.org/single_cell/study/SCP503). To reduce signature enrichment bias due to poor sequencing coverage, samples with less than 10,000 UMIs were excluded from the analysis, and the resulting dataset contained 28 glioma stem cells samples and 6 patient derived GBM samples.

The gene expression profiling was focused on a list of 64 genesets previously described by our group (Lam et al, *Nature Communications*, 2022) which were selected from the MSigDB-7.2 database (https://www.gsea-msigdb.org/gsea/msigdb) on the basis of being informative in glioblastoma and showing a high degree of proteo-transcriptomic correlation. Here, we used a revised list of 64 genesets that differs from the previously published as follows: (i) The pathway “REACTOME_HSP90_CHAPERONE_CYCLE_FOR_STEROID_…” was dropped during the version upgrade to MSigDB-7.4, and (ii) the signature LEIN_ASTROCYTOMA_MARKERS was included as a result of screening for signatures informative of “astrocytic” differentiation. Single sample geneset enrichment analysis (ssGSEA) was performed with the Bioconductor package GSVAv1.44.2.

The Astro-ES axis is a composite score designed to cover the spectrum of cellular differentiation that is found in diffuse glioma, where high values correspond to a “well differentiated” astrocytic phenotype and lower values with an embryonic “poorly differentiated” state. This score is calculated by zero-centering the ssGSEA scores of the LEIN_ASTROCYTOMA_MARKERS and BENPORATH_ES_1 genesets, and then subtracting the latter from the former.

### Estimation of regional cellular density in HAVOC-defined regions

We estimated cell density in profiled regions to complement the interpretation of HAVOC defined regions. To do this WSI data files (.svs format) were loaded on QuPath v0.3.2. For each region of interest, 5 circular sampling areas spanning 100-125 µm^2^ and histologically representative of the region of interest, were delineated, and followed by running Estimate Stain Vectors and Cell Detection Tool with default parameters. The density of each sampling area was computed as follows: number of detections x 100 / area, and finally, the regional cellular density is the average of the cellular density of the 5 sampling areas. Finally, the regional cellular density was categorized according to the following arbitrarily selected reference framework: Ultra-High (>1.2), High (8-1.2), Mid (6-8) and Low (<6).

## Data availability

The mass spectrometry proteomics data of all HAVOC derived regions presented in this manuscript have been deposited to the ProteomeXchange Consortium via the PRIDE partner repository with the dataset identifier PXD037548 (username; password: fy4uPFAi). The data can also be examined directly through our inter-active data portal (Brain Protein Atlas^29^; https://www.brainproteinatlas.org/dash/apps/ad). Labeling legend for mass spectrometry proteomics data can be found at https://bitbucket.org/diamandislabii/havoc.

As described, some of the data used in this publication derived from The Cancer Genome Atlas Program (TCGA) and deposited at the Data Coordinating Center (DCC) for public access [http://cancergenome.nih.gov/]. The RNA-Seq IvyGAP data used are publicly available at Gene Expression Omnibus through GEO series accession number GSE107560. The single-cell are publicly available through the Broad Institute Single-Cell Portal (https://singlecell.broadinstitute.org/single_cell/study/SCP503) and CReSCENT60 (https://crescent.cloud; study ID CRES-P23). Additional proteomic data from different glioblastoma regions were also derived from Lam et al^33^ are also publicly available through the ProteomeXchange Consortium via the PRIDE partner repository with the dataset identifier PXD019381. The H&E slide and ground truth annotations for the metastatic lung carcinoma mouse model was provided directly from the authors and the relevant study^6^.

## Code availability

Code for the 512 dimensional feature vector extractor, feature activation mapping (FAM) and the original trained VGG19 model used in HAVOC is available on Bitbucket (https://bitbucket.org/diamandislabii/faust-feature-vectors-2019 and https://doi.org/10.5281/zenodo.3234829). Python source code and an interactive Colab notebook for running HAVOC and integrating multiple slides into a single analysis are available at https://bitbucket.org/diamandislabii/havoc and https://colab.research.google.com/drive/1Gx7gXTBIBF5iNY0REL7QktM-oAMv4W6S?usp=sharing. HAVOC can also be tested and run directly within a web browser by uploading a .SVS digital WSI on https://www.codido.co

## Notes

### Competing Interest Statement

The authors have declared no competing interest.

### Author Declarations

The University Health Network Research Ethics Board approved the study: REB #17-6193.

