## Supplementary Figures for "HAVOC: Small-scale histomic mapping of biodiversity across entire tumor specimens using deep neural networks"

Supplemental Figure 1

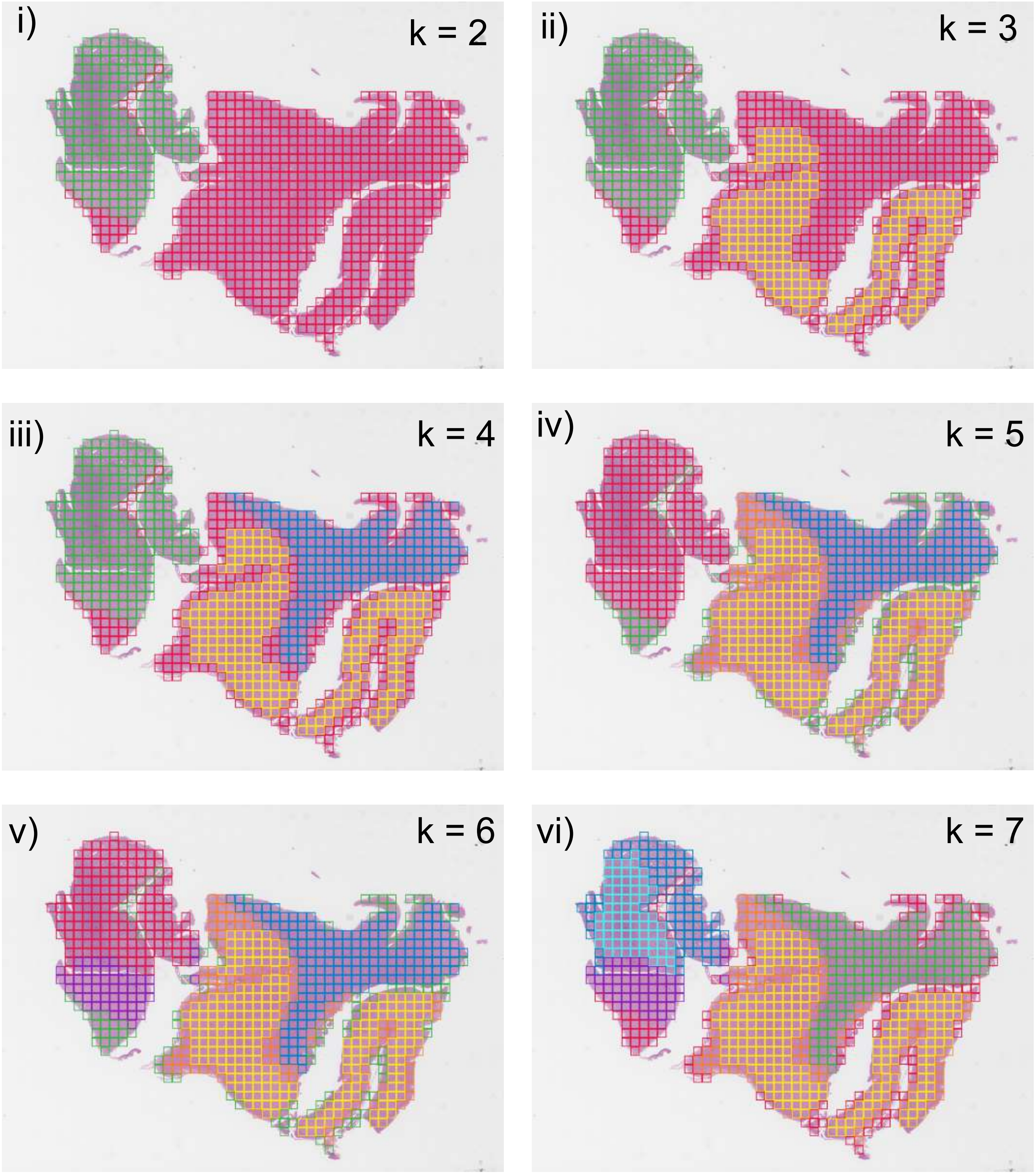

**Supplemental Figure 1 | Sequential image-based clustering solutions resolve different degrees of heterogeneity across WSIs.** Clustering of a diffuse glioma WSI with partition solutions ranging from k=2-7 (i-vi). In most cases, early HAVOC partitions separate large histomorphologic differences such as tumoral vs normal brain tissue (green versus red, k=2). Additional partitions allow for the identification of more subtle histomorphological differences, such as glioma regions with and without edema (cyan vs blue, k=7, respectively).

Supplemental Figure 2

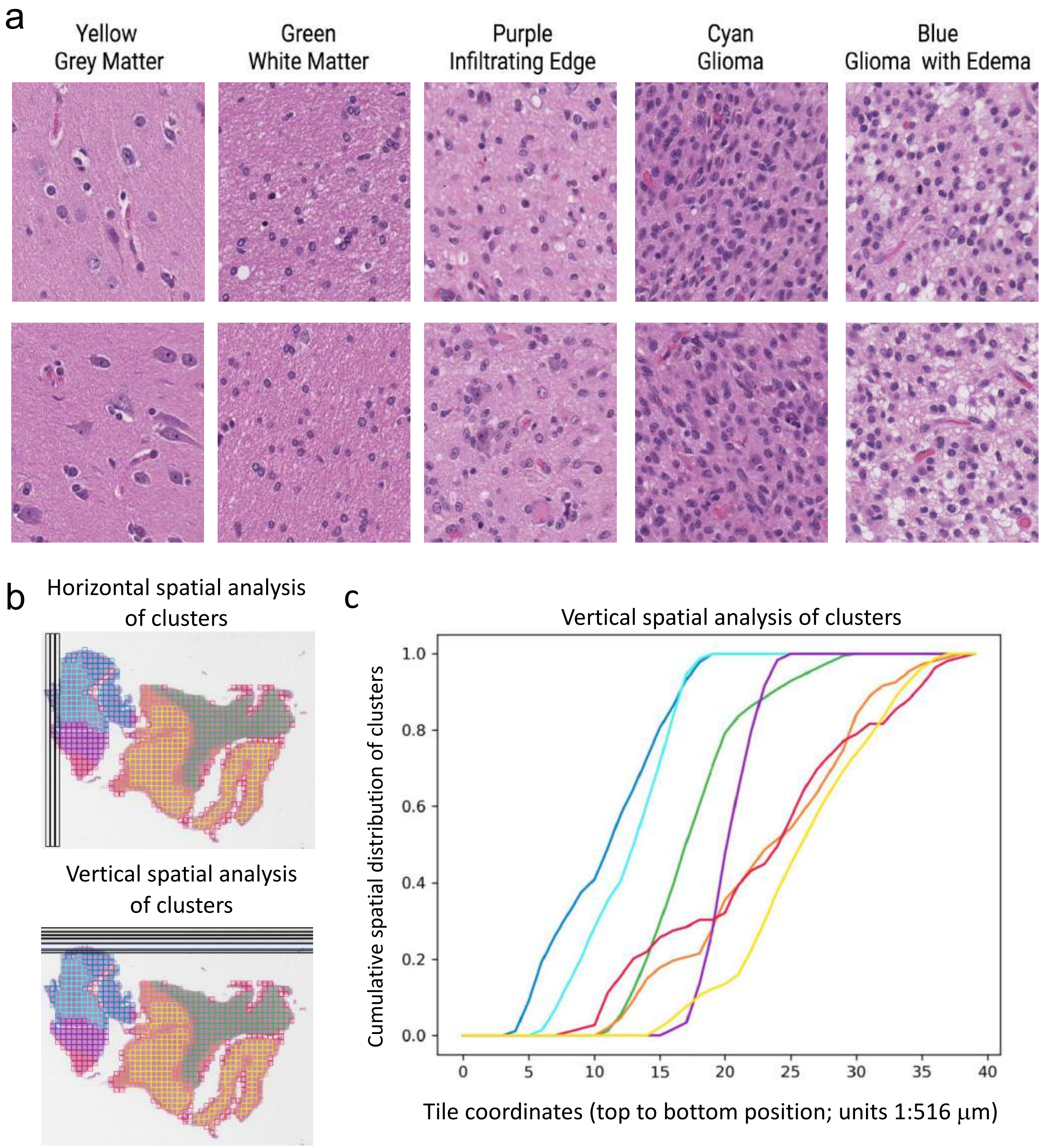

**Supplemental Figure 2 | HAVOC resolution of intra-tumoral niches in diffuse glioma. (a)** Representative image patches from the discrete histomorphologic HAVOC partitions for the slide shown in Fig 1. Regions align with pathologist-defined glioma niches including cellular tumor, infiltrating tumor, white and gray matter. More subtle areas with tumoral edema are also detected. (**b**) Cartoon highlighting how spatial distribution differences of tumor patterns across entire slide were analyzed. (**c**) vertical cumulative distribution plot of the overall fraction of tiles from each HAVOC-defined cluster in Fig 1d across entire WSI highlighting non-random spatial distributions of defined histomorphologies (Kolmogorov–Smirnov statistic of distribution of cellular $\text{tumor}_{\text{cyan}}$ vs infiltrating $\text{tumor}_{\text{purple}}$ = 0.375, p=0.0068).

Supplemental Figure 3

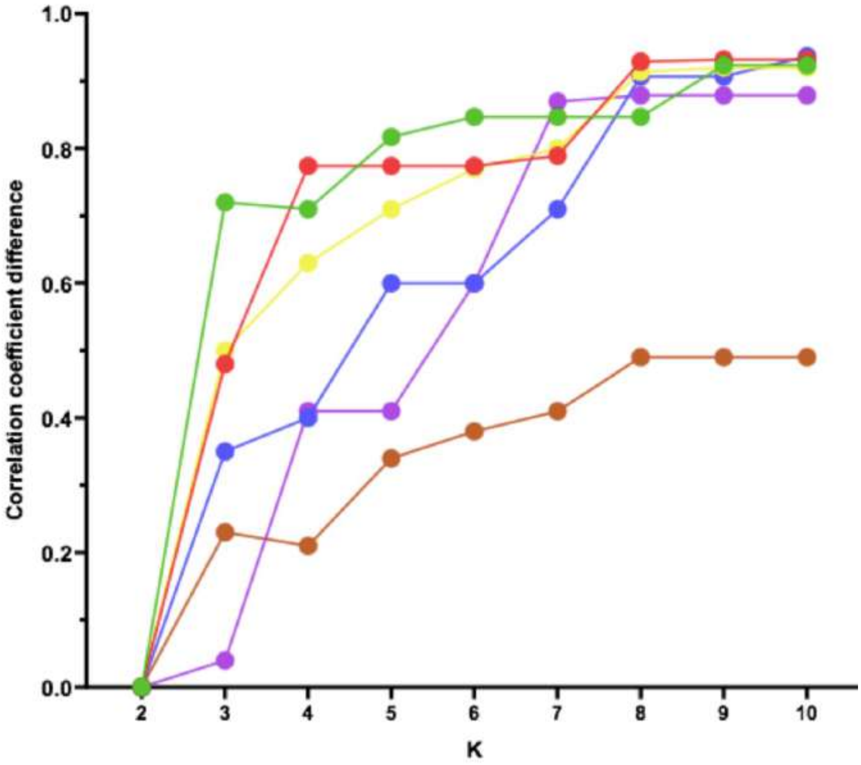

**Supplemental Figure 3 | Highest DLFV *r* between HAVOC-defined partitions across six representative WSI increasing from k=2 to k=8.** In the majority of encountered specimen, irrespective of the initial partition correlation, the degree of pathologist discernible histomorphologic heterogeneity (DLFV *r* ~0.84) reaches saturation after ~6-7 WSI partitions.

Supplemental Figure 4

a

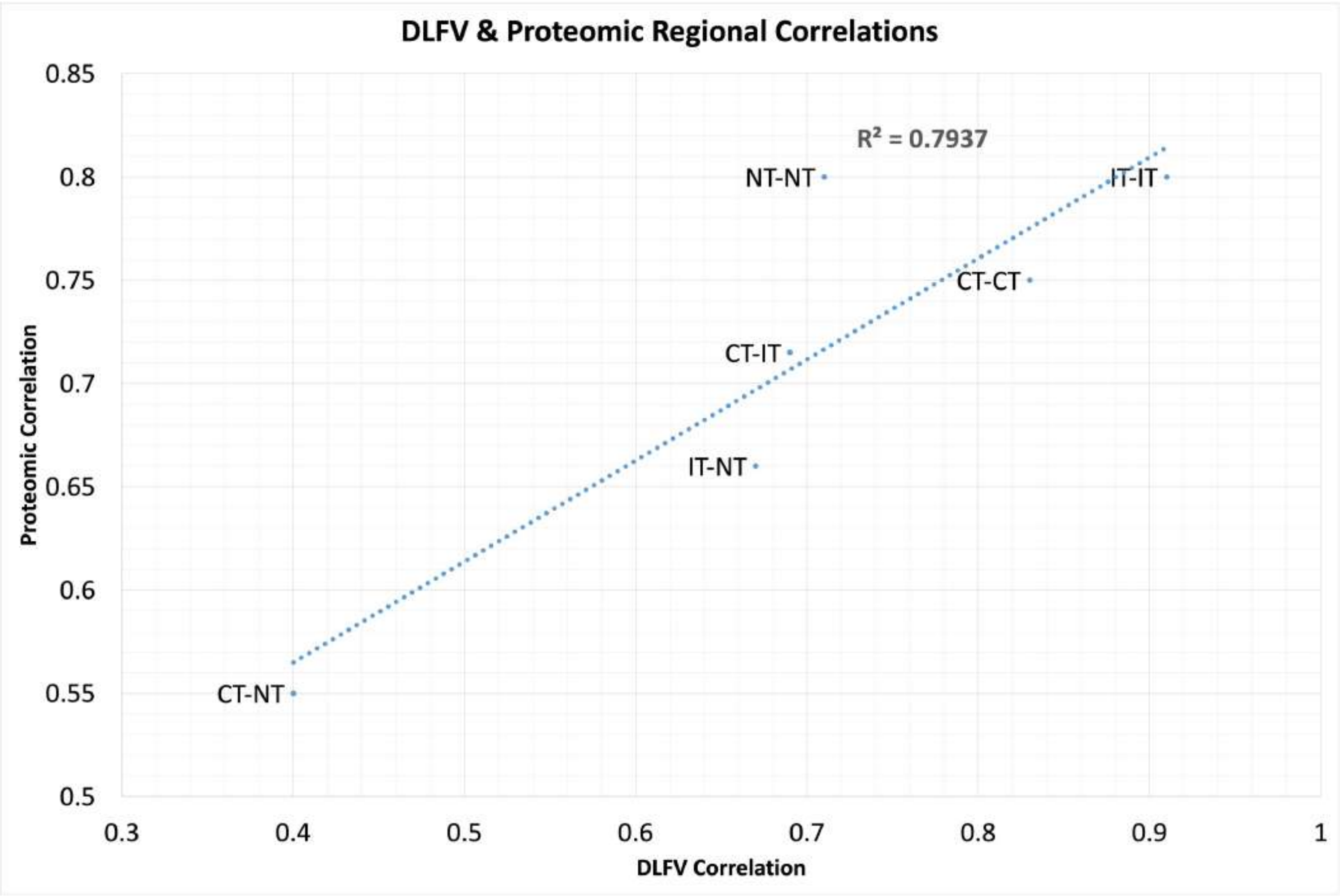


b

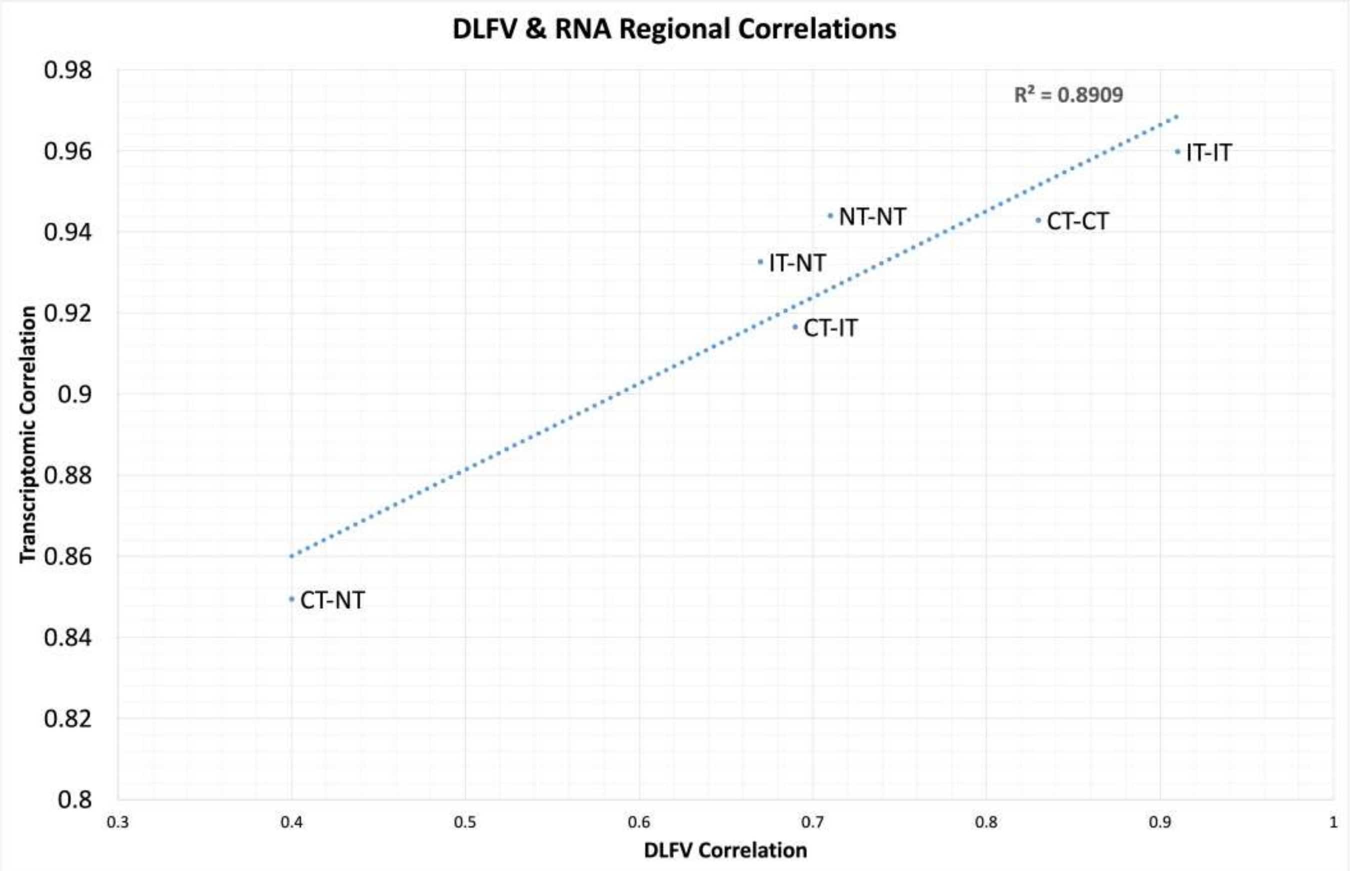

**Supplemental Figure 4 | Molecular expression patterns and DLFVs covary across diffuse glioma niches.** Comparison of region-to-region (a) proteomic and (b) transcriptomic versus DLFV Pearson correlations (*r*) between well-defined glioma niches (leading edge (LE), infiltrating tumor (IT), and cellular tumor (CT)). Both modalities show a strong correlation. Proteomic data retrieved from Lam et al (Nature *Communications*, 2022) and transcriptomic data taken from the Ivy Gap database (Puchalski et al, *Science*, 2018).

### Supplemental Figure 5

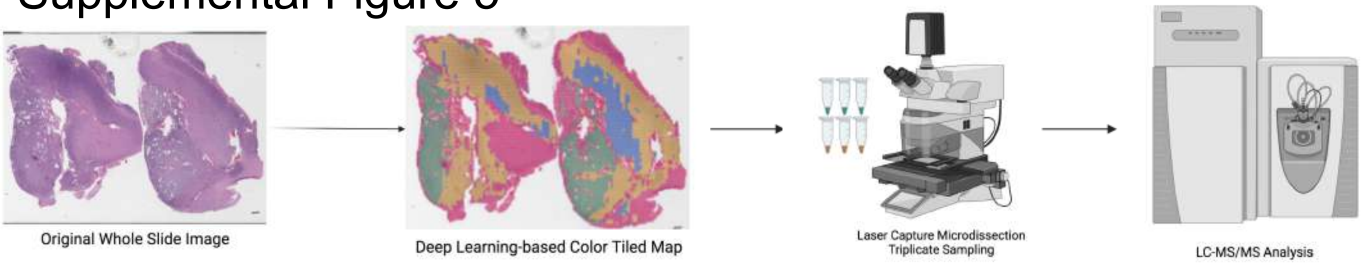

**Supplemental Figure 5 | Schematic of HAVOC profiling workflow.** H&E-stained WSI are assessed by HAVOC to define spatial distribution of intra-tumoral histomorphological differences. HAVOC-defined regions are subsequently isolated with laser-capture microdissection and prepared for LC-MS/MS proteomic profiling.

### Supplemental Figure 6

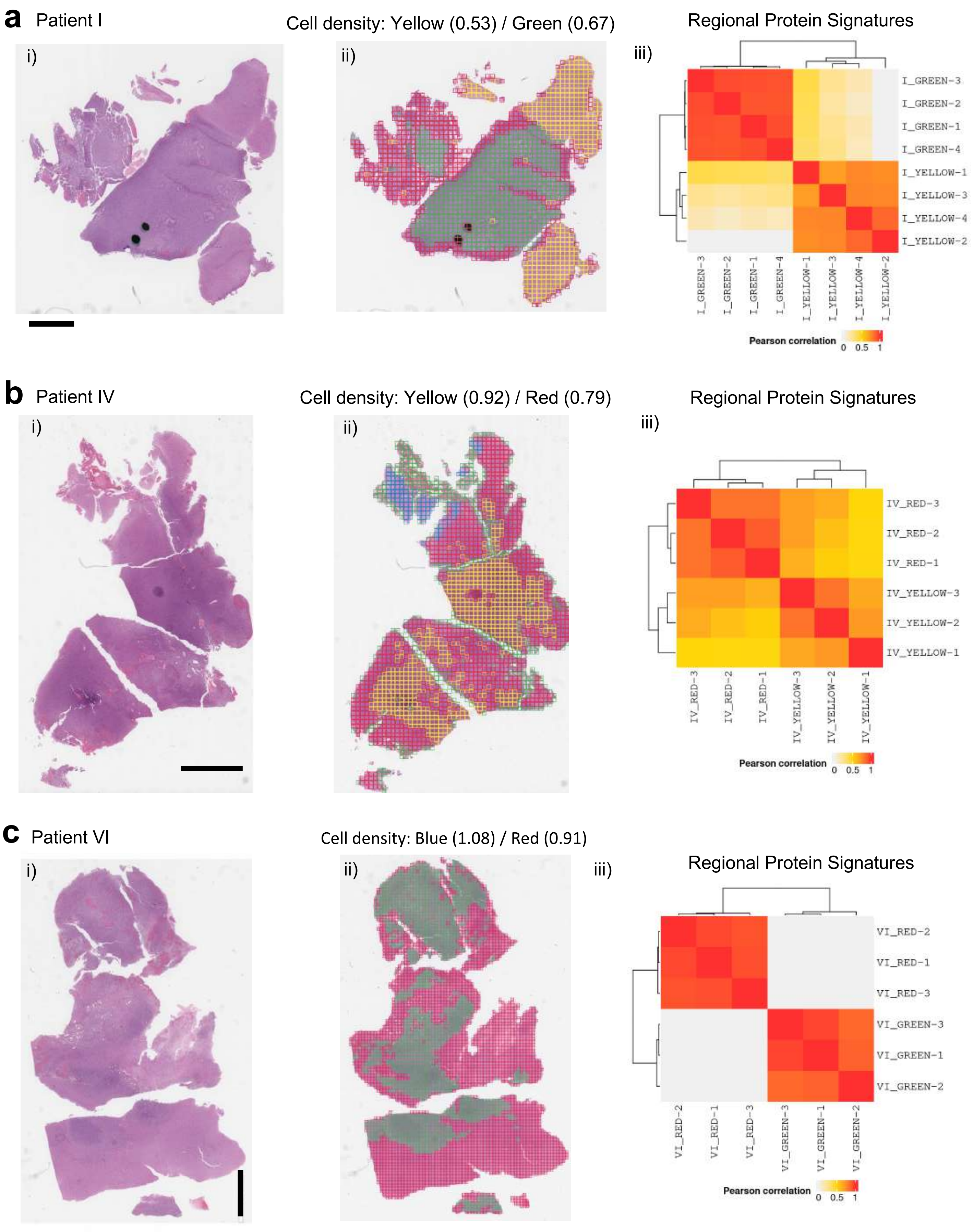

**Supplemental Figure 6 | Mapping of spatial protein-based pathway differences in high grade glioma using HAVOC.** Additional examples of spatial differences in individual cases of diffuse gliomas guided by HAVOC.

#### Supplemental Figure 7

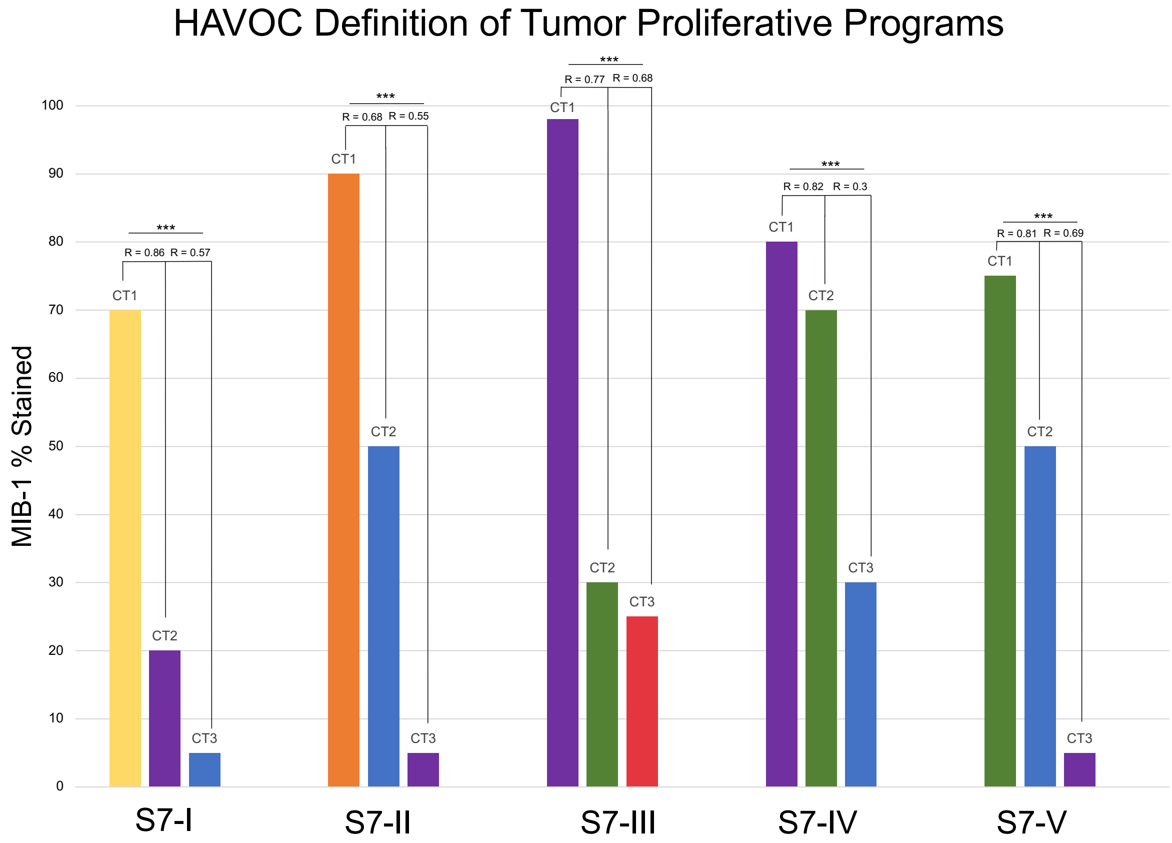


#### Supplemental Figure 8

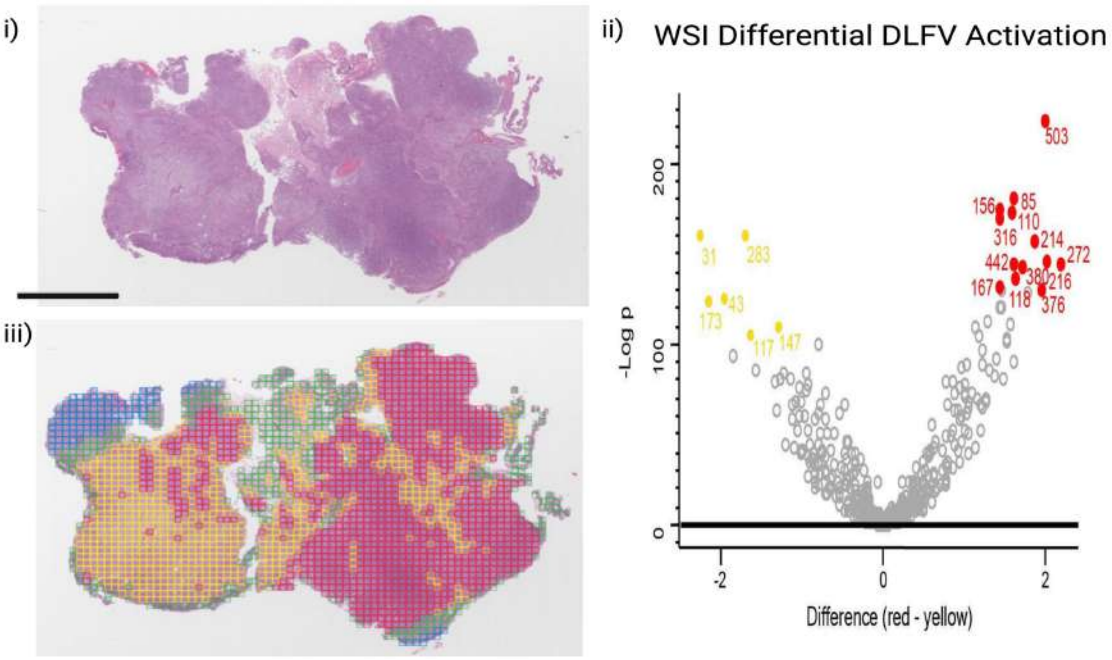

**Supplemental Figure 7 | Histomic heterogeneity HAVOC distances (r) correlate with proliferation differences.** Relationship between estimated proliferation indices (Ki-67) and regional DLFV-generated *r* values across HAVOC partitions. n=4, *** denotes $p<0.0001$.

**Supplemental Figure 8 | Regional variation of deep learning features in a glioblastoma.** Volcano plot highlighting differential DLF activation between the HAVOC-defined partitions. The H&E and HAVOC map from Figure 2 are included here as a reference.

#### Supplemental Figure 9

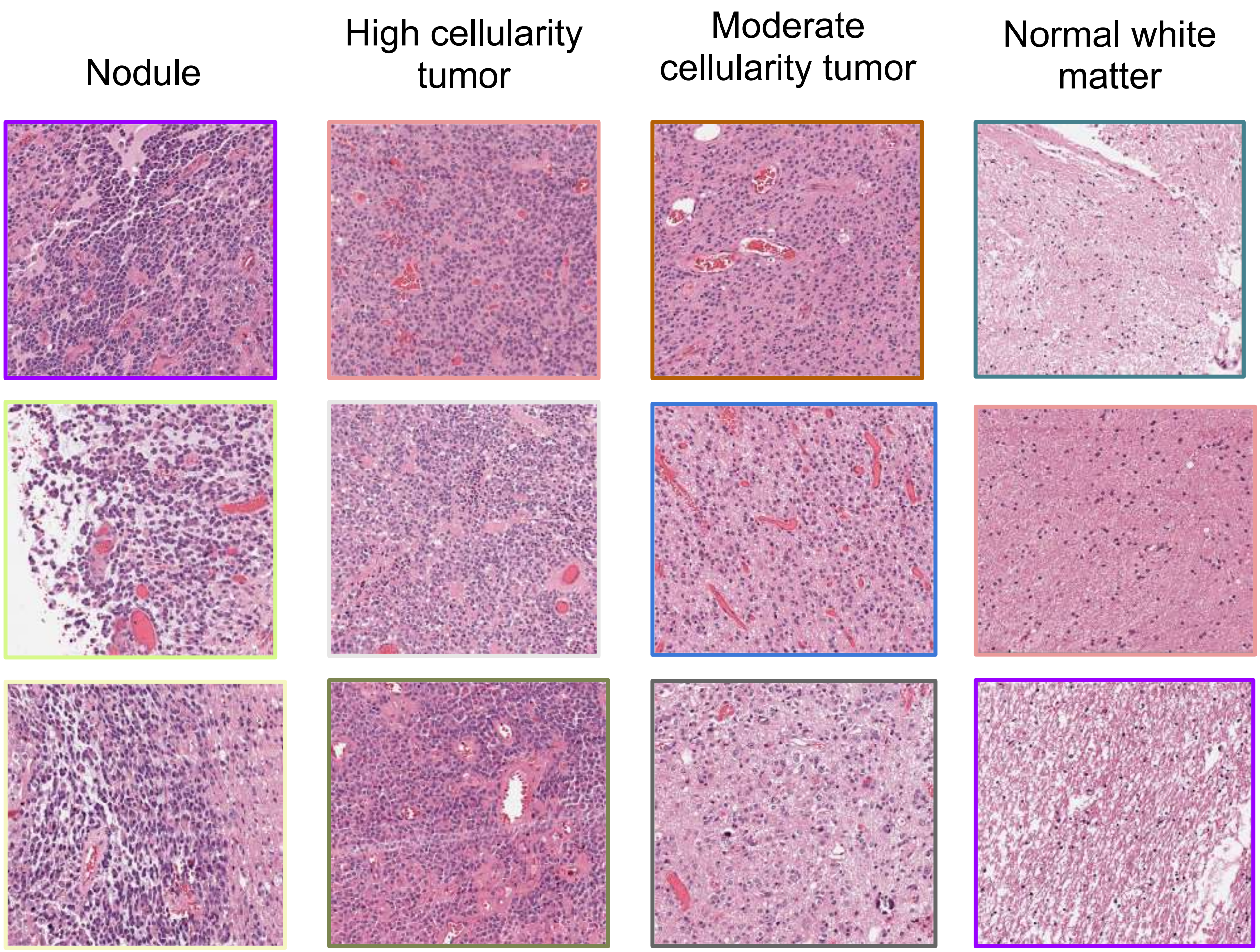


#### Supplemental Figure 10

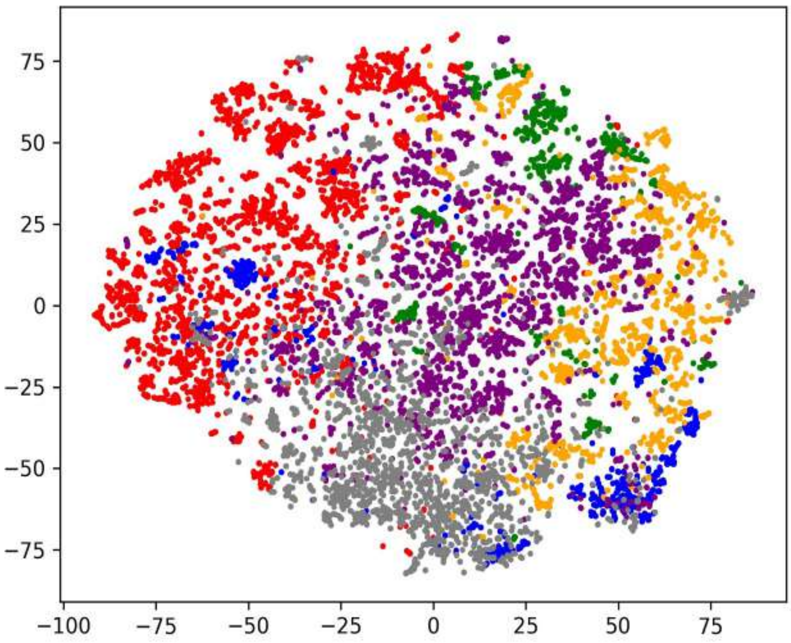

**Supplemental Figure 9 | Representative H&E images patches from different clustered regions from Figure 3.**

**Supplemental Figure 10 | t-SNE plot of all 10,973 0.27 $mm^2$ image patches from the entire tumor in Figure 3.** Coloured tiles correspond to tumor cluster colour IDs indicated in Figure 3c. From right to left, there is a gradient transitioning from hyper-cellular nodules (blue), cellular tumor (orange), purple (moderate 1), green (moderate 2) and non-tumor/low tumor cellularity (red). Gray points highlight non-cellular cluster (hemorrhage/edge tiles).

### Supplemental Figure 11

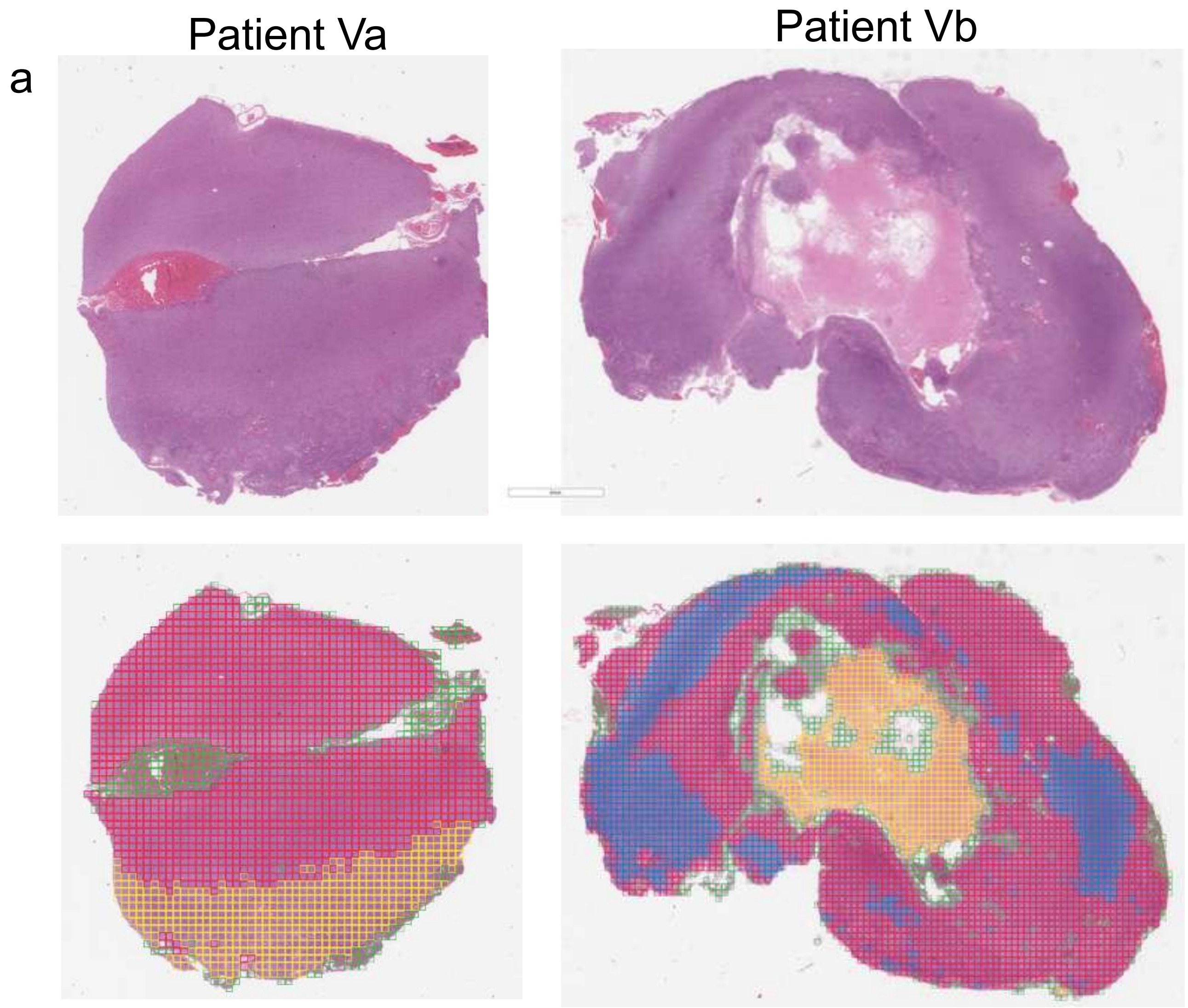


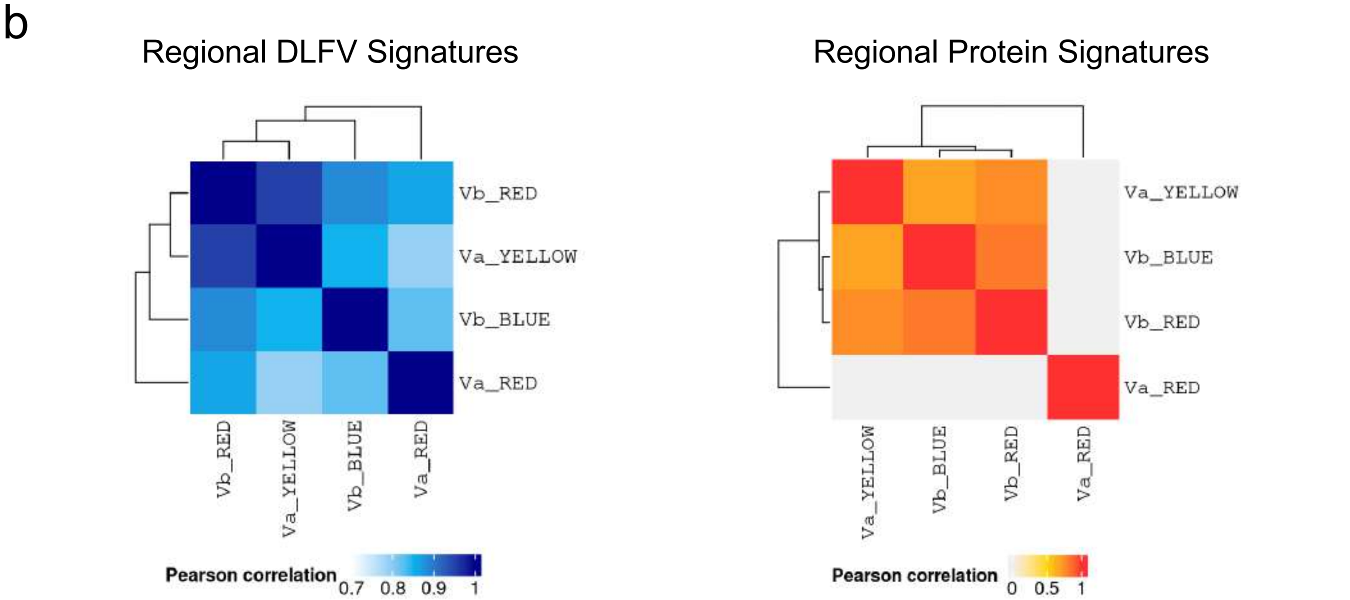

**Supplemental Figure 11 | HAVOC mapping across independent slides align with overall molecular correlations (a)** Another example of a slide pair analyzed concurrently by HAVOC and molecular analysis. **(b)** Integrated DLFV correlation matrices across both slides define a distinct infiltrative compartment and a more cellular tumor region on initial slide. The accompanying slide shows 2 proposed tumor patterns encircling a central necrotic region. These multi-slide maps are in agreement with global patterns of proteomic variations derived from each of the major HAVOC defined partitions.

### Supplemental Figure 12

#### Patient II

| Group | Region (avg. cell density) | |
|---|---|---|
| | IIa | IIb |
| Ultra-high | - | BLUE (1.31) |
| High | GREEN (0.83) | RED (0.94) |
| Mid | YELLOW (0.67) | - |
| Low | - | YELLOW (0.44) |

Block IIa

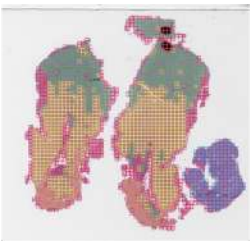

Block IIb

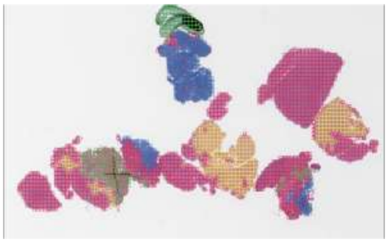

#### Patient III

| Group | Region (avg. cell density) | |
|---|---|---|
| | IIIa | IIIb |
| Ultra-high | - | - |
| High | - | - |
| Mid | YELLOW (0.60) | GREEN (0.72) |
| Low | GREEN (0.46) | RED (0.51) |

Block IIIa

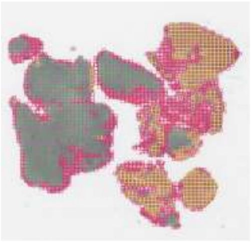

Block IIIb

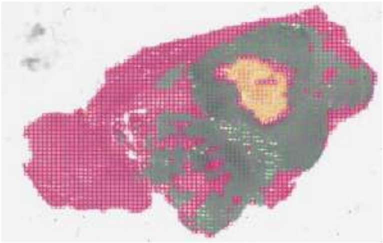

#### Patient V

| Group | Region (avg. cell density) | |
|---|---|---|
| | Va | Vb |
| Ultra-high | - | - |
| High | YELLOW (0.85)<br>RED (0.79) | BLUE (1.09)<br>RED (0.91) |
| Mid | - | - |
| Low | - | - |

Block Va

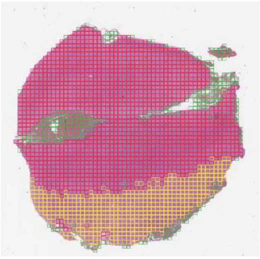

Block Vb

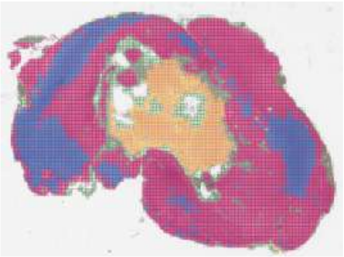

**Supplemental Figure 12 | Regional cell density estimates align with HAVOC mapping across independent slides.** Correlation of cell densities of closely clustered regions further supports the non-random partitioning and grouping of regions by HAVOC.

### Supplemental Figure 13

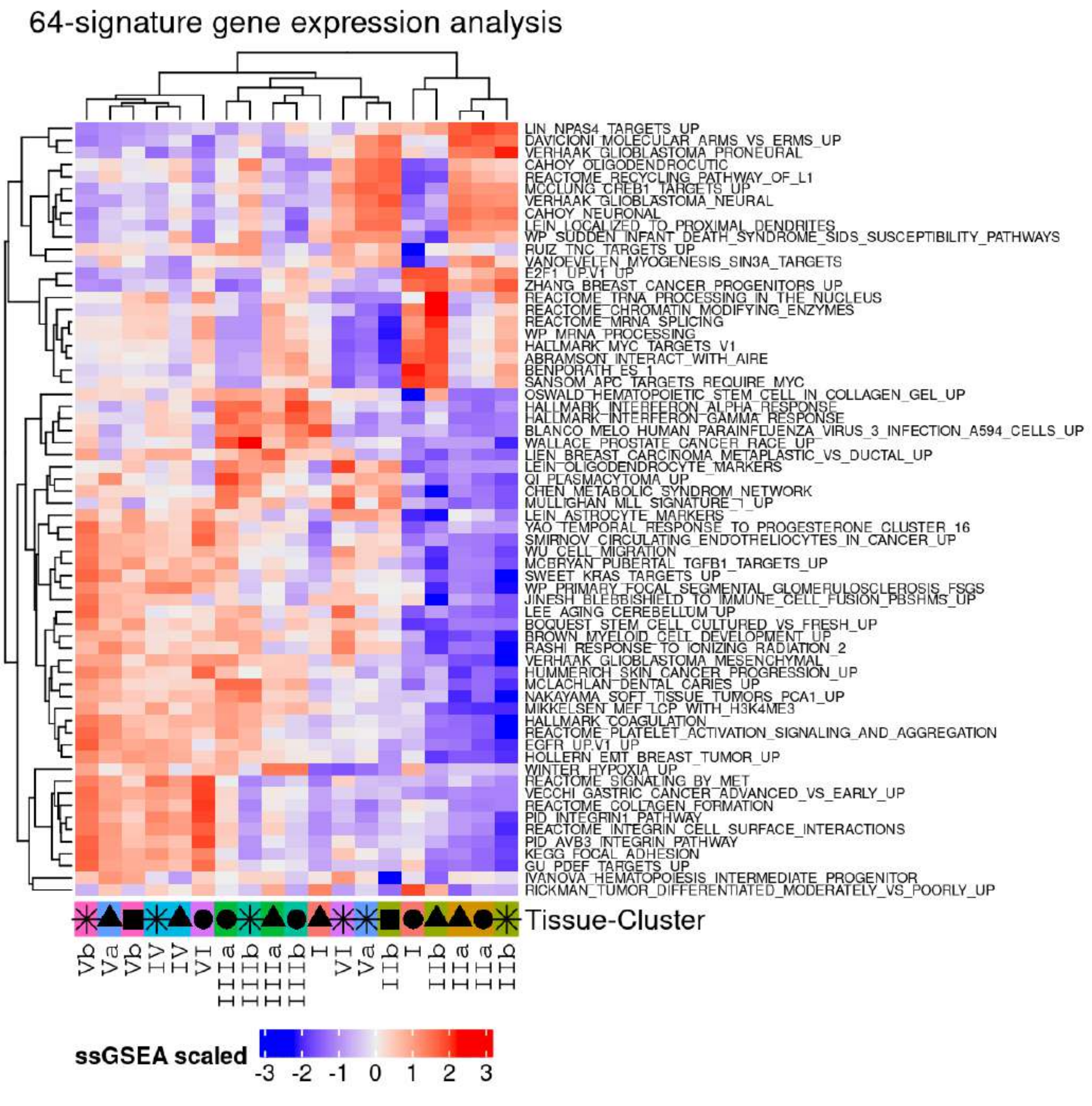

**Supplemental Figure 13 | Hierarchical clustering of proteomics signatures.** Enrichment of 64 pre-selected genesets that were previously deemed informative in GBM (Lam et al, 2022) was calculated by ssGSEA and followed by hierarchical clustering. This unsupervised approach results in samples being mainly grouped by tissue, and with limited intra-sample regional separation.

Supplemental Figure 14

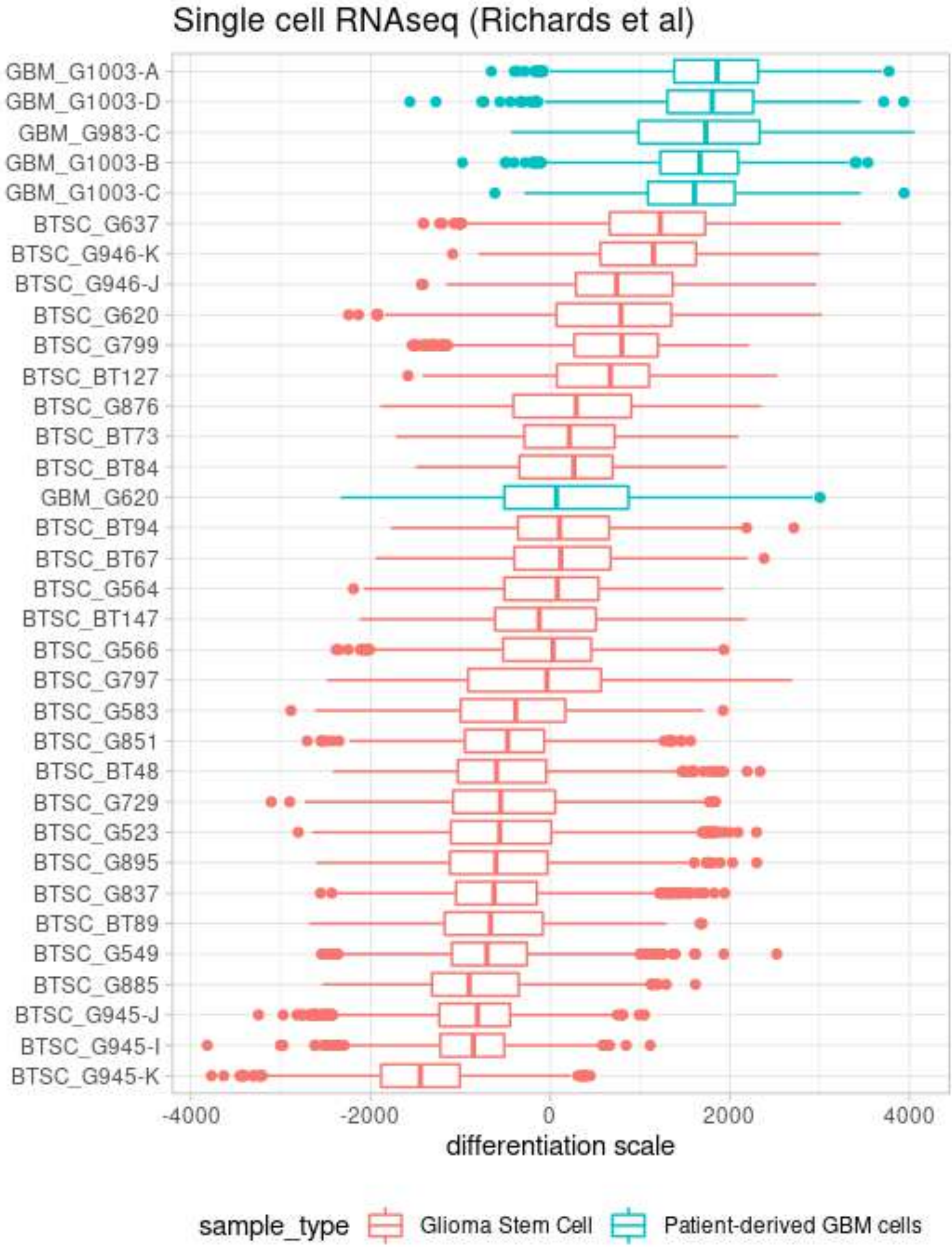

**Supplemental Figure 14 | Varying distributions along the Astro-ES axis at the single-cell level in glioma stem cells (n=28) and patient-derived GBM cells (n=6);** dataset previously published by Richards et al.

Supplemental Figure 15

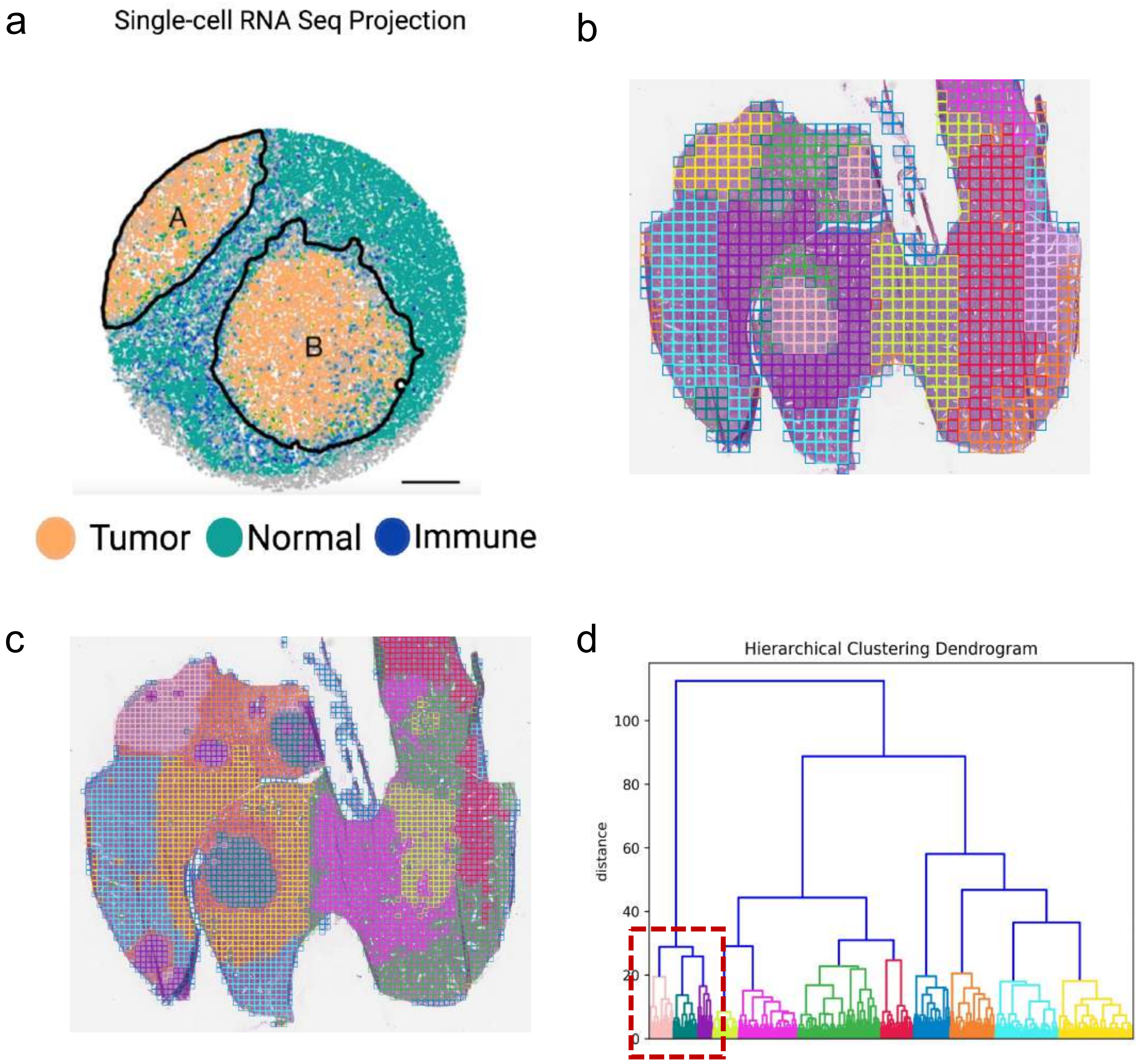

**Supplemental Figure 15 | HAVOC partitioning defines metastatic subclones and peritumoral changes in an experimental lung cancer model. (a)** Spatial RNAseq image taken from Zhao et al. as a reference for ground truth. In addition to lesion segmentation, this approach highlight peritumoral immune infiltrates within the neighbouring liver tissue parenchyma. (**b-c)**. HAVOC analysis at both a tile width of 1024 (panel b) and 512 (panel c) pixels defined five relatively homogeneous tumoral populations. These sub-partitions were relatively stable across the different tile sizes, but smaller tiles sizes provide higher resolution in specific cases in which pattern was predominantly driven by nuclear patterns. (**d)**. Using the silhouette method, the five tumors in panel c. group into two major clusters (highlighted in red).

Supplemental Figure 16

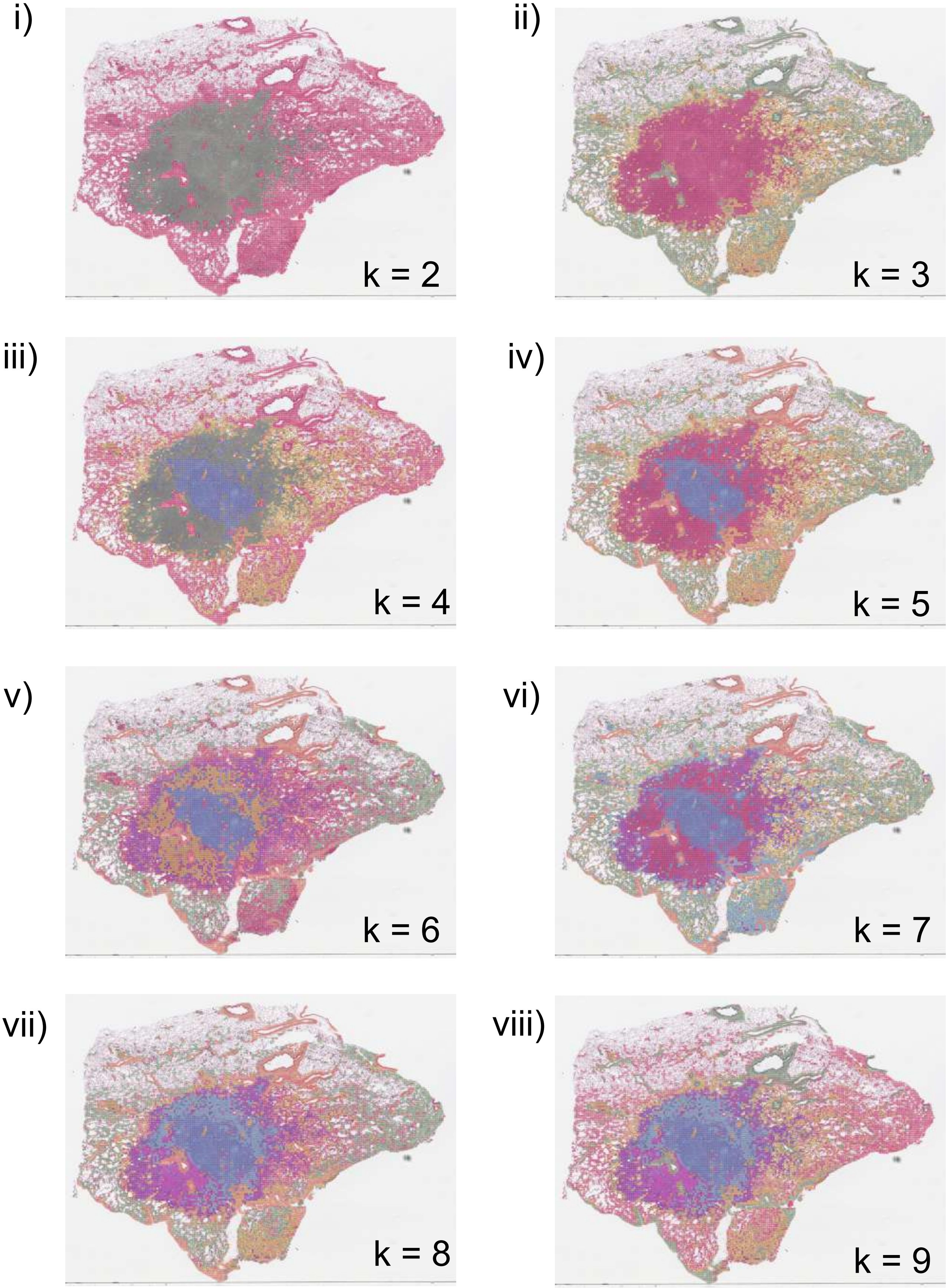

**Supplemental Figure 16 | Sequential image-based clustering solutions resolve different degrees of heterogeneity across WSIs.** Clustering of image patches of a WSI of a lung adenosquamous carcinoma with partition solutions ranging from k=2-9. The major squamoid pattern of morphologic heterogeneity was resolved at k=4 (blue vs green, r = 0.86). Subsequent partitions of the adenosquamous regions showed more minor histomic differences that were not appreciated by human experts as significant (Blue vs Cyan, r = 0.95).

### Supplemental Table 1

**Supplementary Table 1| Clinical and pathological details of high-grade glioma cohort used for HAVOC-based regional profiling**

| Patient ID (Block) | Primary vs Secondary | Age Range | Sex | Location | IDH Status | IDH Method | ATRX (IHC) | p53 (IHC) | MGMT Promoter Methylation (Sequencing) | Diagnosis |
|---|---|---|---|---|---|---|---|---|---|---|
| I (Block 1B) | Primary | 66-70 | Female | Right temporal | WT | IHC | Retained | WT | Positive | Glioblastoma, WHO Grade 4 |
| IIa (Block 3C)<br>IIb (Block 3D) | Primary | 46-50 | Male | Right temporal | WT | Sequencing | Retained | WT | N/A | High grade glioma (WHO grade 3) |
| IIIa (Block 1A)<br>IIIb (Block 1B) | Primary | 56-60 | Male | Right parietal/ frontal | WT | IHC | Retained | Mutant | Negative | Glioblastoma, WHO Grade 4 |
| IV (Block 2F) | Primary | 51-55 | Male | Right parietal/ frontal | WT | IHC | Retained | WT | Negative | Glioblastoma, WHO Grade 4 |
| Va (Block 2A)<br>Vb (Block 2D) | Primary | 56-60 | Female | Right frontal | WT | IHC | Retained | WT | Positive | Glioblastoma, WHO Grade 4 |
| VI (Block 2C) | Primary | 56-60 | Male | Left parietal/ occipital | WT | IHC | Retained | WT | N/A | Glioblastoma, WHO Grade 4 |

### Supplemental Table 2

**Supplementary Table 2| Clinical and pathological details of additional representative cases analyzed by HAVOC.** For convivence, Patient ID represents figure (e.g. “F1”) and panel (e.g. C) where this case was introduced. N/A denotes not available

| Patient ID | Primary vs Secondary | Age Range | Sex | Location | IDH Status | IDH Method | ATRX (IHC) | p53 (IHC) | MGMT Promoter Methylation (Sequencing) | Diagnosis |
|---|---|---|---|---|---|---|---|---|---|---|
| F1C (Block 2E) | Primary | 51-55 | M | Right temporal | WT | Sequencing | Retained | WT | Negative | Anaplastic Astrocytoma, IDH-wildtype, WHO Grade 3 |
| F2A (Block 2A) | Primary | 66-70 | Female | Right temporal | WT + BRAFV600 E Subclone | Sequencing | Retained | WT | Negative | Glioblastoma, WHO Grade 4 |
| F2B (Block 2A) | Primary | 66-70 | Male | Left frontal | WT | IHC | Retained | N/A | Positive | Glioblastoma, WHO Grade 4 |
| F2H, SF7-II (Block 2B) | Primary | 56-60 | Male | Left temporal | WT | IHC | Retained | WT | Negative | Glioblastoma, WHO Grade 4 |
| F3B (Blocks 1A-1L) | Recurrent | 51-55 | Male | Left frontal | Mutant | IHC | Retained | WT | N/A | Anaplastic Oligodendroglioma. IDH-mutated, 1p19q codeleted, WHO Grade 3 |
| SF7-I (Block 1A) | Primary | 56-60 | Male | Left parietal | WT | IHC | Retained | N/A | Negative | Glioblastoma, WHO Grade 4 |
| SF7-III (Block 2C) | Primary | 61-65 | Male | Right frontal | WT | IHC | Retained | Mutant | Positive | Glioblastoma, WHO Grade 4 |
| SF7-IV (Block 2F) | Primary | 56-60 | Male | Left frontal | Mutant | IHC | Retained | WT | N/A | Anaplastic Oligodendroglioma. IDH-mutated, 1p19q codeleted, WHO Grade 3 |
| SF7-V (Block 2A) | Primary | 66-70 | Male | Right frontal | WT | IHC | Retained | Mutant | Negative | Glioblastoma, WHO Grade 4 |
